# Association of polygenic risk scores for depression, anxiety, and neuroticism with lower urinary tract symptoms in women

**DOI:** 10.1101/2025.08.04.25332951

**Authors:** Zayn Rajan, Christina Dardani, Kimberley Burrows, Oliver Bastiani, Marcus J Drake, Carol Joinson

## Abstract

**Background:** Observational studies have found that depression and anxiety are prospectively associated with lower urinary tract symptoms (LUTS), but these studies are limited by potential environmental confounding and measurement error. Using genetic liability for psychiatric traits offers an alternative approach.

**Objectives:** Examine the association of genetic liability for depression, anxiety, and neuroticism with LUTS in women.

**Methods:** Data: i) Avon Longitudinal Study of Parents and Children (ALSPAC) Mothers Cohort. ii) GWAS (genome-wide association study) summary data on psychiatric traits.

Exposures: Polygenic risk scores (PRS) for depression, anxiety, and neuroticism.

Outcomes: LUTS - assessed in 2002-04 (mean [sd] age = 40.3 [4.6]) and 2011-12 (49.7 [4.5]) using validated questionnaires.

Statistical analysis: Logistic regression adjusted for age and population structure.

**Key findings and limitations:** The neuroticism PRS was associated with nocturia at both time points [odds ratio and 95% confidence intervals=1.24 (1.10,1.40) and 1.21 (1.07,1.37)], and with any UI, any urgency, and mixed UI (2002-04 only) [1.14 (1.05,1.23), 1.12 (1.02,1.23), and 1.23 (1.07,1.43), respectively]. The depression PRS was associated with nocturia [1.33 (1.17,1.50) and 1.24 (1.09,1.40)] and any UI [1.09 (1.01,1.18); 1.11 (1.02,1.20)] at both timepoints, and with any urgency (2011-12 only) [1.13 (1.03,1.24)]. The anxiety PRS was associated with mixed UI (2002-04 only) [1.20 (1.03,1.39)].

PRS may not capture all genetic liability and cannot account for pleiotropic effects. The ALSPAC cohort includes only parous women and is predominantly affluent and of European ancestry.

**Conclusions and clinical implications:** Genetic liability for depression, anxiety and neuroticism was associated with LUTS in women, suggesting psychiatric factors may contribute to the aetiology of LUTS.

**Patient summary:** In this study we looked at whether parous women with a high genetic risk of mental health problems were more likely to have urinary symptoms. We found various associations, with the strongest evidence found for genetic liability to depression and neuroticism increasing the risk of nocturia (waking during the night to pass urine). Our findings suggest that psychiatric factors could be important in understanding urinary symptoms, and more research is needed to find out if this these associations apply to other demographics.

## Introduction

Urinary incontinence (UI) is a lower urinary tract symptom (LUTS) which around 10% of women experience at least weekly [1]. The most prevalent subtype is stress UI (prevalence: 10-39%), which is urine leakage during exertion, such as when coughing or laughing [1].

Urgency UI (1-7%), urine leakage following a strong and sudden need to urinate, is less common [1]. Mixed UI (7.5-25%) is a combination of stress UI and urgency UI [1]. In this study, “any UI” is used to capture individuals experiencing either stress UI, urgency UI, or mixed UI. Another common type of LUTS is nocturia - waking during the night to void (prevalence: 11-14%) [2].

Comorbidity of UI with depression and anxiety is well-established in cross-sectional studies [3]. A systematic review found cross-sectional associations of both depression and anxiety with nocturia [4]. An observational study in women with overactive bladder (OAB) found a positive correlation of daily urinary urgency scores with both depression and anxiety [5].

Cross-sectional studies cannot provide insights into the direction of associations between depression/anxiety and LUTS. Some prospective studies that examined associations between LUTS and depression/anxiety have provided evidence of bidirectional associations [6,7]. A prospective cohort study in parous women (mean age=41.0) found that depression was associated with subsequent LUTS (any UI, mixed UI and urgency), whilst anxiety was only associated with subsequent nocturia [8]. The study found little evidence that LUTS were associated with subsequent depression or anxiety; only stress UI with subsequent depression [8]. A cohort study of older women (mean age=50.3) found evidence that depressive symptoms were associated with subsequent LUTS [9]. Neuroticism, one of the ‘big five’ domains of personality characterised by a disposition to experience negative affect including anxiety, and depression [10], was prospectively associated with UI in a cohort of women (mean age=46.8) [11]. Evidence that mental health problems are associated with subsequent LUTS warrants further investigation because this contradicts the common assumption that poor mental health is a consequence, not a potential cause, of LUTS.

Although prospective studies can provide evidence of the direction of the association between depression/anxiety and LUTS, these studies are limited by unmeasured and residual confounding [12]. Measurement error is another limitation of prospective studies that rely on self-report questionnaires to assess psychiatric traits [13]. An alternative approach is the use of polygenic risk scores (PRSs) as estimates of genetic liability for complex traits including depression, anxiety and neuroticism [14]. Since genetic variants are randomly inherited at meiosis and fixed at conception, PRSs should not be associated with environmental confounders at a population level and any identified associations with outcomes of interest should not be a result of reverse causation.

This study, based on data from a population-based cohort of middle-aged parous women in the UK, uses PRSs to estimate the associations of genetic liability for depression, anxiety, and neuroticism with LUTS in women. We assessed LUTS using validated questionnaires at two timepoints and we hypothesise that higher PRSs for depression, anxiety, and neuroticism will be associated with an increase in the odds of LUTS.

## Participants and methods

### Polygenic Risk Scores (PRS)

PRS enable the estimation of an individual’s genetic liability to a complex trait (see section 3.1 of the Supplement). PRSs for depression, anxiety, and neuroticism were calculated for all participants with sufficient genotype data.

### Study population

The women included in this study are from the Avon Longitudinal Study of Parents and Children (ALSPAC) [15–17]. This is a population-based, prospective cohort study which followed pregnant women (and their newborns) resident in Avon, with a total sample size of 15,447 pregnancies. (Further detail in Section 1 of the Supplement).

Figure 1 illustrates the analysis samples for the study. Differing sample sizes are a result of questionnaire completion rates and attrition between the 2 timepoints. Associations of PRS for depression, anxiety, and neuroticism with LUTS were examined in Sample 3 and Sample 4 for 2002-04 and 2011-12 respectively.

**Figure 1:**
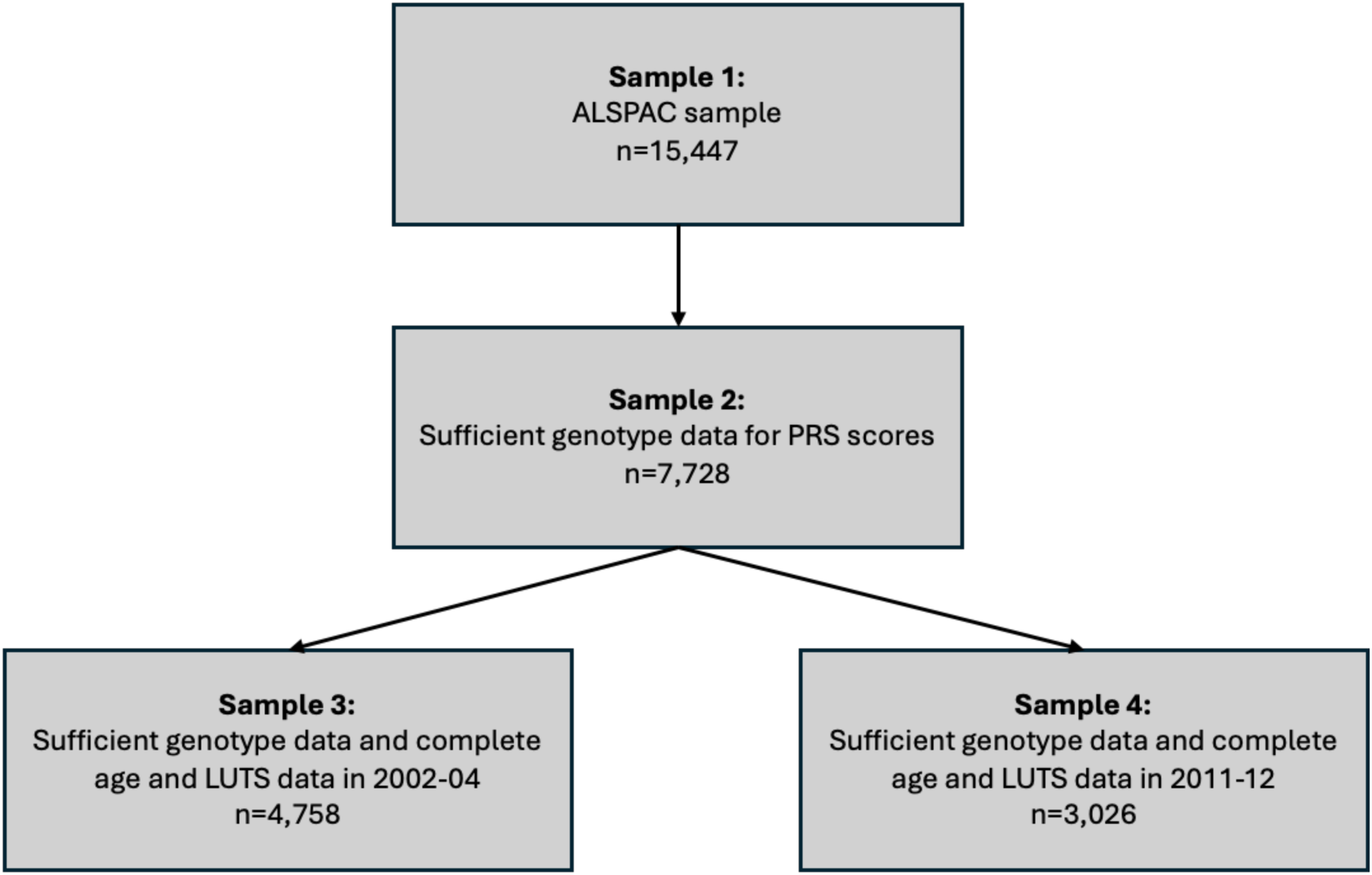
Flow diagram of analysis samples. LUTS = Lower Urinary Tract Symptoms

### Discovery sample: Genome-wide association studies (GWASs)

An explanation of GWASs is provided in Section 2 of the Supplement.

GWAS summary data were used to extract genetic variants associated with depression [18], anxiety [19], and neuroticism [20] (further detail in Supplementary Table 2.1).

### Target sample

ALSPAC mothers were genotyped using the Illumina human660W-quad array at Centre National de Genotypage (CNG) and genotypes were called with Illumina GenomeStudio. PLINK v1.07 was used to carry out quality control measures on an initial set of 10,015 participants and 557,124 directly genotyped SNPs.

After quality control and excluding participants who had withdrawn consent, sufficient genetic data for PRS derivation were available for 7,728 participants.

### PRS Estimation

PRSs were calculated using PLINK v.1.9, applying the method described by the Psychiatric Genomics Consortium (PGC) [21]. This approach is consistent with current guidelines and is the most widely used method for deriving PRS of psychiatric traits. (See section 3.2 of the Supplement).

We assessed the performance of depression and anxiety PRSs by testing their associations with validated questionnaire-based measures of depression and anxiety respectively, which were administered to the mothers of the ALSPAC birth cohort at 18 weeks of gestation (see Section 3.4 of the Supplement).

### Measures – Lower urinary tract symptoms (LUTS)

Participants completed the Bristol Female LUTS (BFLUTS) in 2002-04 and the International Consultation on Incontinence Questionnaire on Female LUTS (ICIQ-FLUTS) in 2011-12 [22].

Stress UI was defined by the question “*Does urine leak when you are physically active, exert yourself, cough or sneeze?*”. Urgency UI was defined by the questions “*Does urine leak before you can get to the toilet?*” and “*Do you have a sudden need to rush to the toilet to urinate?*” Participants who met the criteria for both stress UI and urgency UI were defined as mixed UI cases. Participants who were defined in any of the three UI categories (stress, urgency, or mixed) were classified as having any UI. Any urgency was defined by the question “*Do you have a sudden need to rush to the toilet to urinate?*”. Responses to the questions were recorded using a 5-point Likert scale with the following options: *“Never”, “Occasionally”, “Sometimes”, “Most of the time”, “All of the time”*. Participants who responded with at least *“Sometimes”* were defined as cases.

Nocturia was assessed by the question “*What is the number of times you urinate throughout the night?*” Cases were defined as those waking to urinate two or more times.

## Statistical analysis

The association of the PRSs for depression, anxiety, and neuroticism with LUTS were examined using logistic regression. All models were adjusted for age (at the time of LUTS questionnaire) and the first 10 genetic principal components (PCs) in ALSPAC. (See Section 3.3. of the Supplement)

Associations were separately examined between 3 psychiatric traits and 6 LUTS phenotypes, at 2 timepoints, resulting in 36 tests. False discovery rate (FDR) adjusted p-values were used to control type 1 error across the tests (<5%) [23].

The primary analysis was based on PRS corresponding to p<0.05, which is consistent with previous ALSPAC studies [24].

A sensitivity analysis excluded participants with organic causes of LUTS (see Section 5 of the Supplement).

### Evidence Appraisal

We appraised our findings in terms of consistency of the estimates and respective confidence intervals across multiple PRS p-value thresholds (p<0.5 to p<5×10^-8^) for SNP inclusion. On this basis we could infer whether the identified primary associations (even those passing FDR) were not a result of chance. This is in line with existing guidelines suggesting that evidence appraisal in terms of statistical significance could be misleading and should be avoided [25].

## Results

Table 1 presents the demographics and prevalences of LUTS within each analysis sample. The proportions of women categorised as manual social class and those with the lowest level of educational attainment were lower in the more restricted samples. The prevalence of LUTS was similar across the different samples and was higher at the later (2011-12), compared with the earlier (2002-04), timepoint.

**Table 1:** Demographics and prevalences of LUTS (lower urinary tract symptoms) across analysis samples. Question completion rates may differ within the same sample.

|  | **Sample 1:** | **Sample 2:** | **Sample 3:** | **Sample 4:** |
| --- | --- | --- | --- | --- |
|  |  | *Genotype data* |  |  |
|  | *ALSPAC*<br>*sample*<br>*(n=15,447)* | *(n=7,728)* | *Genotype data*<br>*and complete*<br>*age and LUTS*<br>*data in 2002-04*<br>*(n=4,758)* | *Genotype data*<br>*and complete*<br>*age and LUTS*<br>*data in 2011-12*<br>*(n=3,026)* |
| Age in 2002-04: |  |  |  |  |
| Median (IQR) | 40.0 (37,43) | 40.0 (37,43) | 40.0 (37,43) | 41.0 (38,44) |
| Range | 27-56 | 27-56 | 27-56 | 27-54 |
| Age in 2011-12: |  |  |  |  |
| Median (IQR) | 49.0 (47,53) | 50.0 (47,53) | 50.0 (47,53) | 50.0(47,53) |
| Range | 36-64 | 36-64 | 36-64 | 36-64 |
| Maternal Social<br>Class: |  |  |  |  |
| Manual<br>(ref: non-manual) | 1,983 (19.9%) | 1,013 (17.3%) | 578 (14.6%) | 316 (11.7%) |
| Maternal education: |  |  |  |  |
| CSE/Vocational |  |  |  |  |
| O-level | 3,706 (30.1%) | 1,751 (25.2%) | 869 (19.2%) | 440 (14.7%) |
| (ref: A-level or<br>greater) | 4,262 (34.6%) | 2,400 (34.6%) | 1,549 (34.3%) | 986 (32.9%) |
| **2002-04**<br>**prevalence:** |  |  |  |  |
| Stress UI | 771 (10.3%) | 501 (10.4%) | 495 (10.4%) | 283 (10.2%) |
| Urgency UI | 107 (1.4%) | 70 (1.5%) | 70 (1.5%) | 46 (1.6%) |
| Mixed UI | 327 (4.4%) | 188 (3.9%) | 187 (3.9%) | 101 (3.6%) |
| Nocturia | 469 (6.3%) | 282 (5.9%) | 278 (5.8%) | 139 (5.0%) |
| Any UI | 1,205 (16.1%) | 759 (15.8%) | 752 (15.8%) | 430 (15.4%) |
| Any urgency | 858 (11.5%) | 508 (10.6%) | 504 (10.6%) | 267 (9.6%) |
| **2011-12**<br>**prevalence:** |  |  |  |  |
| Stress UI | 614 (13.9%) | 409 (13.5%) | 371 (13.4%) | 409 (13.5%) |
| Urgency UI | 117 (2.6%) | 87 (2.9%) | 80 (2.9%) | 87 (2.9%) |
| Mixed UI | 392 (8.9%) | 256 (8.5%) | 234 (8.4%) | 256 (8.5%) |
| Nocturia | 399 (9.0%) | 273 (9.0%) | 245 (8.8%) | 273 (9.0%) |
| Any UI | 1,123 (25.4%) | 752 (24.9%) | 685 (24.7%) | 752 (24.9%) |
| Any urgency | 791 (17.9%) | 542 (17.9%) | 490 (17.7%) | 542 (17.9%) |

Figure 2 shows odds ratios, 95% confidence intervals (CI), and p-values for the association of PRSs for anxiety, depression and neuroticism with LUTS at the p<0.05 SNP-inclusion threshold. The results are also presented in Supplementary Table 4.1.

**Figure 2:**
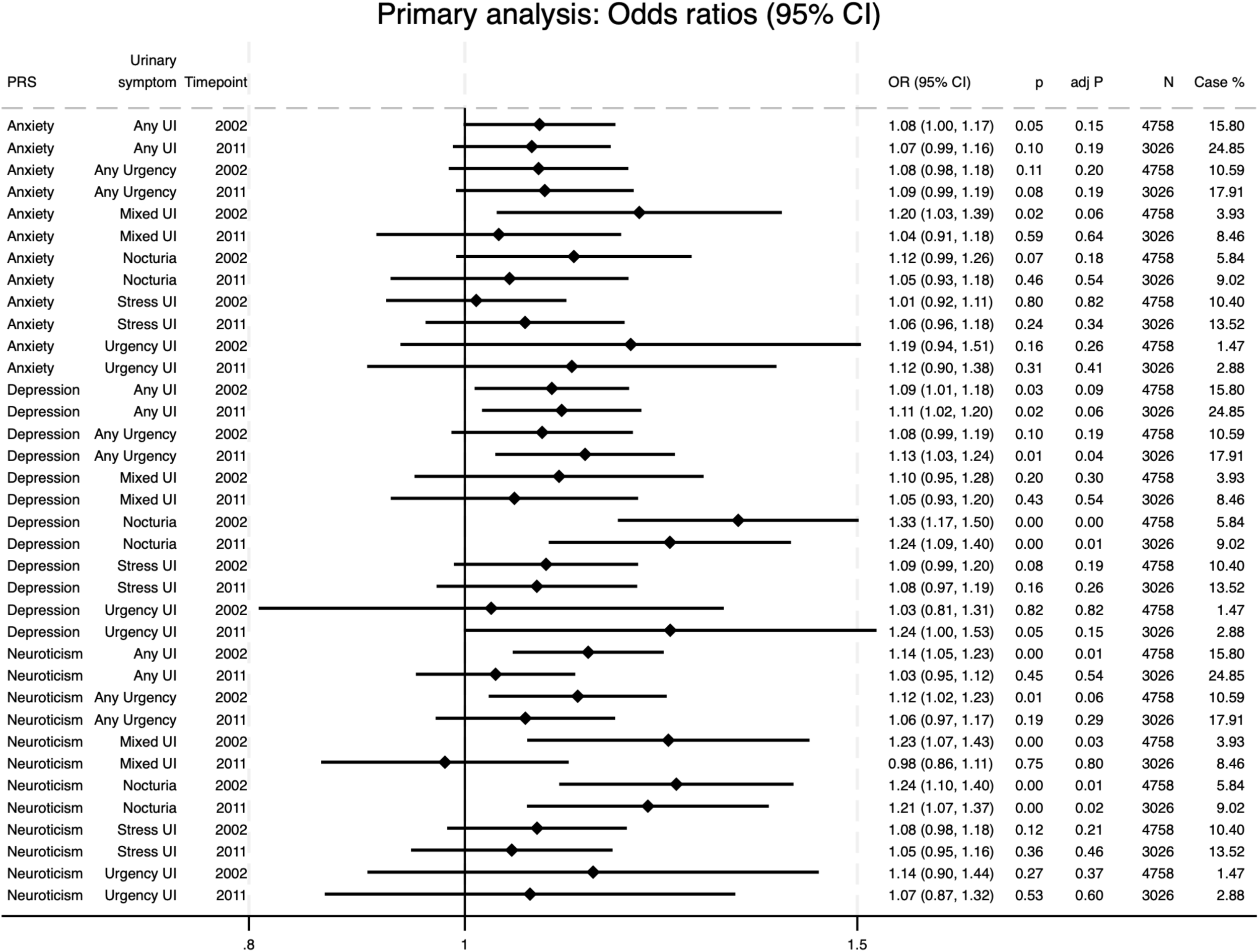
Associations of polygenic risk scores for depression, anxiety, and neuroticism (at the p<0.05 SNP-inclusion significance threshold) with lower urinary tract symptoms. All models are adjusted for age and the first 10 principal components in ALSPAC. The figure illustrates the odds ratios and 95% confidence intervals, unadjusted p-values, adjusted p-values controlling for FDR, the number of participants in each model, and the percentage of these participants who were positive cases for the urinary symptom.

The odds ratios for the associations between the PRSs and LUTS were all above one, except for neuroticism and mixed UI (2011-12). The greatest number of associations were found for the neuroticism and depression PRSs, which were strongly associated with nocturia at both timepoints (2002-4 and 2011-12). The neuroticism PRS was also associated with any UI, any urgency, and mixed UI (2002-4 only). The depression PRS was also associated with any UI at both timepoints and any urgency at the second timepoint (there was weak evidence of an association at the first timepoint). The anxiety PRS was only associated with mixed UI (2002-4 only). It is notable that many of these associations survived FDR adjustment.

Specifically, neuroticism and depression PRSs with nocturia (both timepoints), depression PRS with any urgency (second timepoint), and neuroticism PRS with any UI and mixed UI (first timepoint).

Weak evidence was found for associations of the anxiety PRS with any UI (both timepoints), nocturia (first timepoint), and any urgency (second timepoint). The depression PRS was weakly associated with stress UI (first timepoint), and urgency UI (second timepoint).

The pattern of results did not change in the sensitivity analysis (Supplementary Figure 5.1).

A similar pattern of associations was seen across the other SNP-inclusion thresholds (Supplementary Figures 6.1-6.12).

## Discussion

We found evidence that genetic liability to depression, anxiety, and neuroticism is associated with LUTS in middle-aged women, implying that women with these psychiatric traits may have an increased risk of developing LUTS. Our study took a novel approach by using PRS as opposed to questionnaire-based measures of mental health. The strongest evidence we found was for associations of depression and neuroticism PRSs with nocturia.

### Strengths

A strength of this study is the use of data from a large, prospective, community-based cohort, and the use of validated questionnaires to assess LUTS, including UI subtypes. Although voiding diaries are the ideal diagnostic tool for evaluating nocturia [26], this was not feasible for a large cohort study. However, studies commonly rely on self-reported nocturia, which can provide valuable insight into the psychology of symptom reporting and the subjective experience of nocturia.

The use of PRS captures genetic liability to complex traits, which cannot be influenced by environmental factors after conception that could confound the association between psychiatric traits and LUTS. This is an advantage over using questionnaire-based measures of mental health, which are prone to confounding and measurement error.

We tested 36 associations and reported FDR-adjusted p-values. While many remained significant after adjustment, some did not. Rather than relying solely on p-value thresholds (e.g., p < 0.05), we interpreted results based on effect estimates, p-values, and confidence intervals, in line with current recommendations [27]. We also examined consistency across multiple SNP-inclusion thresholds (Supplementary Figures 6.1–6.12) to ensure findings were not artifacts of any single threshold.

### Limitations

PRS explain a portion of heritable risk, but they may not capture all genetic influence relating to psychiatric traits, particularly contributions from rare genetic variants [28]. Furthermore, the use of PRS cannot account for horizontal pleiotropic effects, whereby a genetic variant may influence both psychiatric traits and LUTS independently, leading to a non-causal association between them [29]. Alignment of our findings with those from observational studies, which have different sources of bias, support the possibility of our findings reflecting underlying biological relationships [30].

Psychological concerns around PRS include false reassurance in those with low scores, and increased anxiety or overtreatment in those with high scores [31]. Ethical contentions include potential genetic discrimination and the risk of exacerbating health inequalities due to Eurocentric GWAS data [31].

Neuroticism and depression showed the strongest associations with LUTS. The neuroticism GWAS used a continuous score for the phenotype, offering greater statistical power and capturing the trait’s continuous variation, unlike the binary case definitions used for depression and anxiety [32].

There was little evidence for associations with stress or urgency UI subtypes. The low prevalence of urgency UI (1.4%), aligning with other studies (1-7%) [1], may have resulted in limited statistical power. Further studies, using validated questionnaires for LUTS, are needed to determine if depression, anxiety and neuroticism are differentially associated with UI subtypes.

The ALSPAC cohort includes only parous women and is predominantly affluent and of European ancestry, limiting the generalisability of our findings to nulliparous women, other ethnicities, or more socioeconomically deprived groups. The GWAS samples used to derive the PRS were also based on individuals of European ancestry, and since PRS performance is population-specific [33], applicability to other backgrounds is uncertain.

### Comparison with previous ALSPAC study

Our findings using depression PRS broadly agree with an earlier study of the ALSPAC cohort associating self-reported depression with subsequent LUTS, but with limited evidence of associations between anxiety and LUTS (except nocturia) [8]. The smaller sample size of the GWAS for anxiety (N=83,566), compared with depression (N=1,349,887) and neuroticism (N=329,821), may explain the weaker associations with anxiety in our study. Few previous longitudinal studies examined both depression and anxiety in relation to LUTS [6,7], highlighting the need for further research on their potentially distinct effects.

### Potential mechanisms

Associations with the neuroticism PRS (and to a lesser extent, the anxiety PRS) were stronger at the earlier timepoint (median age 40) than later (median age 50), potentially due to reduced statistical power at the later timepoint. Alternatively, psychological factors may play a larger role earlier in life, while age-related factors including wider use of medications and menopausal physiological changes (e.g. reduced bladder capacity) may dominate later [34].

Similarly, the depression PRS was more strongly associated with nocturia earlier, but showed a stronger link with urgency later, possibly reflecting its higher prevalence at the later timepoint (17.9% vs. 10.6%).

A Swedish cohort study found evidence that genetic factors in common between depressive mood disorders and urgency UI or mixed UI were entirely shared with neuroticism [35], implying that genetic variants for neuroticism may be driving associations between depression and certain subtypes of UI.

The bladder-gut-brain axis is a conceptual framework to understand the co-occurrence of urological and psychiatric conditions, with chronic stress affecting both emotional regulation and organ function [36]. Neuroticism, by increasing stress sensitivity, may contribute to lower urinary tract symptoms (LUTS).

Mood disorders and LUTS are both associated with elevated inflammation, which may underpin a bidirectional relationship. [37]. Depressed patients (and those with high levels of neuroticism) may lack the typical nocturnal elevation in antidiuretic hormone (ADH) levels, resulting in a higher volume of urine at night [38]. Reductions in monoamines believed to be involved in depression, such as serotonin and noradrenaline, are associated with bladder symptoms including urinary frequency in animal models [39].

### Clinical implications

Our findings suggest that genetic liability to neuroticism, depression, and to a lesser extent anxiety, is associated with an increased risk of LUTS in women, indicating a potential shared aetiology. However, further research is needed to assess causality, clarify underlying mechanisms, and determine generalisability beyond a population of parous women of predominantly white European ancestry. Assessing the potential treatment effect of mental health interventions on LUTS is another important area of focus for future studies. Poor adherence to OAB treatments [40] may be partly explained by coexisting mental health issues; thus, screening for these conditions may improve treatment adherence and outcomes.

## Data Availability

Access to ALSPAC research data must be requested using the formal procedures (outlined at: https://www.bristol.ac.uk/alspac/researchers/access/)
and is subject to eligibility, compliance against the ALSPAC funders terms and conditions and University of
Bristol policies and procedures.

## Acknowledgements

We are extremely grateful to all the families who took part in this study, the midwives for their help in recruiting them, and the whole ALSPAC team, which includes interviewers, computer and laboratory technicians, clerical workers, research scientists, volunteers, managers, receptionists and nurses.

## Funding

The UK Medical Research Council and Wellcome (Grant ref: 217065/Z/19/Z) and the University of Bristol provide core support for ALSPAC. Genomewide genotyping data was generated by Sample Logistics and Genotyping Facilities at Wellcome Sanger Institute and LabCorp (Laboratory Corporation of America) using support from 23andMe.

This research was funded in whole, or in part, by the Wellcome Trust [Grant number: 218495/Z/19/Z]. For the purpose of Open Access, the author has applied a CC BY public copyright licence to any Author Accepted Manuscript version arising from this submission.

This work is supported by funding from the Medical Research Council (grant ref: MR/V033581/1: Mental Health and Incontinence).

This publication is the work of the authors and they will serve as guarantors for the contents of this paper. A comprehensive list of grants funding is available on the ALSPAC website (http://www.bristol.ac.uk/alspac/external/documents/grant-acknowledgements.pdf). The funder had no role in the study design; collection, analysis and interpretation of data; writing of the report; and the decision to submit the article for publication.

## Notes

### Competing Interest Statement

The authors have declared no competing interest.

### Author Declarations

Ethical approval for the study was obtained from the ALSPAC Ethics and Law Committee and the Local Research Ethics Committees. Consent for biological samples has been collected in accordance with the Human Tissue Act (2004). Informed consent for the use of data collected via questionnaires and clinics was obtained from participants following the recommendations of the ALSPAC Ethics and Law Committee at the time. Specific details on the ethics committees and institutional review boards are available here: http://www.bristol.ac.uk/alspac/researchers/research-ethics/

