## Supplement for "Association of polygenic risk scores for depression, anxiety, and neuroticism with lower urinary tract symptoms in women"

### 1. Study population

The participants of this study are from the Avon Longitudinal Study of Parents and Children (ALSPAC), which followed pregnant women (and their newborns) resident in Avon, with expected delivery dates between April 1991 and December 1992.
14,541 pregnancies were initially enrolled. Following an attempt to bolster the sample 7 years into the study, 906 new pregnancies were added to the study resulting in a total sample size of 15,447 pregnancies. Detailed information can be found at the cohort website (www.bristol.ac.uk/alspacwww.bristol.ac.uk/alspac), including a searchable data dictionary and variable search tool: http://www.bristol.ac.uk/alspac/researchers/our-data/.

Ethical approval for the study was obtained from the ALSPAC Ethics and Law Committee and the Local Research Ethics Committees. Consent for biological samples has been collected in accordance with the Human Tissue Act (2004). Informed consent for the use of data collected via questionnaires and clinics was obtained from participants following the recommendations of the ALSPAC Ethics and Law Committee at the time. Specific details on the ethics committees and institutional review boards are available here: http://www.bristol.ac.uk/alspac/researchers/research-ethics/

### 2. Genome-wide association studies (GWAS)

GWASs test for associations between genetic variants and phenotypes in a population, in a hypothesis-free manner [1]. Typically, GWASs focus on single nucleotide polymorphisms (SNPs), which refer to a variation in a single base pair of the DNA and are common in the population (frequency >1%). Associations with the phenotypes of interest are assessed using linear (continuous phenotype) or logistic (binary phenotype) regression models and reported in blocks of SNPs (loci), i.e., SNPs that are co-inherited and correlated referred to as being in high linkage disequilibrium (LD). Loci that pass a p-value threshold ≤5*10-08 are typically considered to be associated with the phenotype of interest at genome-wide significance.

It is increasingly being recognised that for complex traits such as autism, schizophrenia and IQ, the genome-wide significant loci are of small effect size, explain only a small proportion of the genetic and phenotypic variance, and they span across the genome [2]. In fact, complex traits seem to be influenced by a large number of variants bellow the genome-wide threshold and of small effect size- these traits are referred to as polygenic [3]. In polygenic traits, liability to the trait is conceptualised to be normally distributed to the population and individuals are expected to express the phenotype after a threshold of genetic and environmental risk as well as chance, has been exceeded [4,5]. Individuals close but below the threshold are expected to present some sub-phenotypic features (the liability-threshold model of inheritance is visualised in Figure below) [6].

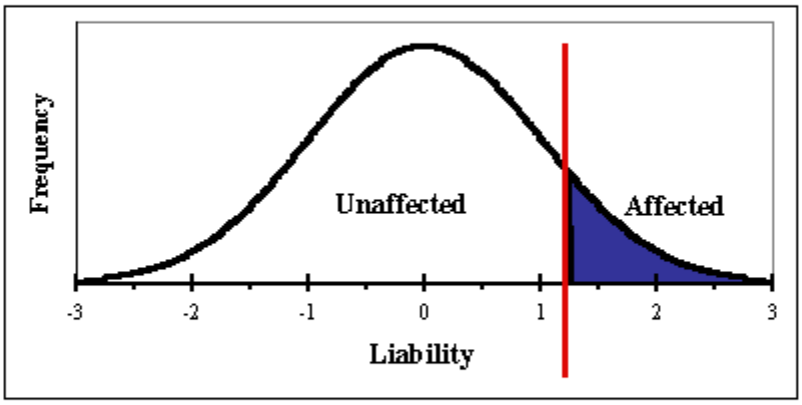

Previous GWASs were used to identify genetic variants associated with depression [7], anxiety [8], and neuroticism [9]. These are detailed in Table 2.1.

#### Table 2.1

| Psychiatric trait | Method | Participants |
|---|---|---|
| Depression | GWAS meta-analysis of 6 datasets | N(case) = 371,184<br><br>N(control) = 978,703 |
| Anxiety | UK Biobank GWAS | N(case) = 25,453<br><br>N(control) = 58,113 |
| Neuroticism | UK Biobank GWAS | N = 329,821 |

### 3. Polygenic Risk Scores (PRS)

#### 3.1 PRS Explanation

PRS approaches enable the estimation of an individual's underlying genetic liability to a complex trait [10]. PRS require individual level genotype data and are calculated as the sum of the individual's risk alleles, weighted by the effect sizes of each variant identified in the

GWAS of the trait. PRS can be calculated for a subset of variants based on a p-value specified threshold, e.g., 5*10-08. Considering that variance in most polygenic traits is unlikely to be explained by genome-wide significant variants, PRS using subsets of variants meeting more relaxed association thresholds e.g., p≤0.05 might capture more variance in the phenotype-which is the case for polygenic traits.

An illustrative example is provided below. Three SNPs are included in the PRS calculation:

| SNP | Effect size | Genotype | Number of risk alleles (G) | Effect size x No. of risk alleles |
|---|---|---|---|---|
| rs1 | 0.10 | AG | 1 | 0.10 x 1 = 0.10 |
| rs2 | 0.20 | GG | 2 | 0.20 x 2 = 0.40 |
| rs3 | 0.05 | AA | 0 | 0.05 x 0 = 0.00 |

PRS = 0.10 + 0.40 + 0.00 = 0.50

#### 3.2 PRS Estimation

We removed single nucleotide polymorphisms (SNPs) with mismatching alleles between the discovery and target dataset. The Major Histocompatibility Complex (MHC) region was removed (25–34 Mb), except for one SNP representing the strongest signal within the region. Using ALSPAC data as the reference panel, SNPs were LD clumped with an $r^2$ of 0.25 and a physical distance threshold of 500 kB. The optimal P-value threshold for PRS is dependent on discovery and target sample sizes, as well as SNP inclusion parameters (for example, $r^2$). PRS for each participant were calculated across 13 P-value thresholds ($P < 5.0 \times 10{-}8$ to $P < 0.5$), standardized by subtracting the mean and dividing by the standard deviation.

#### 3.3 Adjusting for PCs

Principal Components (PCs) are derived from principal component analysis and summarise the genetic structure of individuals into a few dimensions. PCs are included in analysis models to account for population stratification [11]. This occurs when there are distinct subgroups within the population which differ genetically and may also have different environmental exposures. For example, people with different ancestries having different allele frequencies and different disease risks. This can lead to spurious associations if not corrected for.

#### 3.4 Association between PRSs and questionnaire-based depression/anxiety

To assess the performance of depression and anxiety PRSs, we fitted regression models with PRS as the predictor, and questionnaire-based measures of depression and anxiety as the outcome. These models were fitted on all ALSPAC mothers with available genotype data and questionnaire-based measures of depression and anxiety, so that PRS performance could be assessed on the largest possible sample.

**Depression:**

Depression was measured at 18 weeks' gestation using the clinically validated Edinburgh Postnatal Depression Scale (EPDS) [12]. A binary indicator was derived from the EPDS, with a score of 12 or more indicating the presence of a depressive disorder [12]. Thirteen logistic regression models were fitted, one for each SNP-inclusion p-value threshold. All odds ratios (ORs) and associated 95% confidence intervals (CIs) were above 1, indicating strong associations between depression PRSs and questionnaire-based depression.

#### Figure 3.1

Associations between observed depression (Edinburgh post-natal depression score) and depression PRSs at each p-value threshold

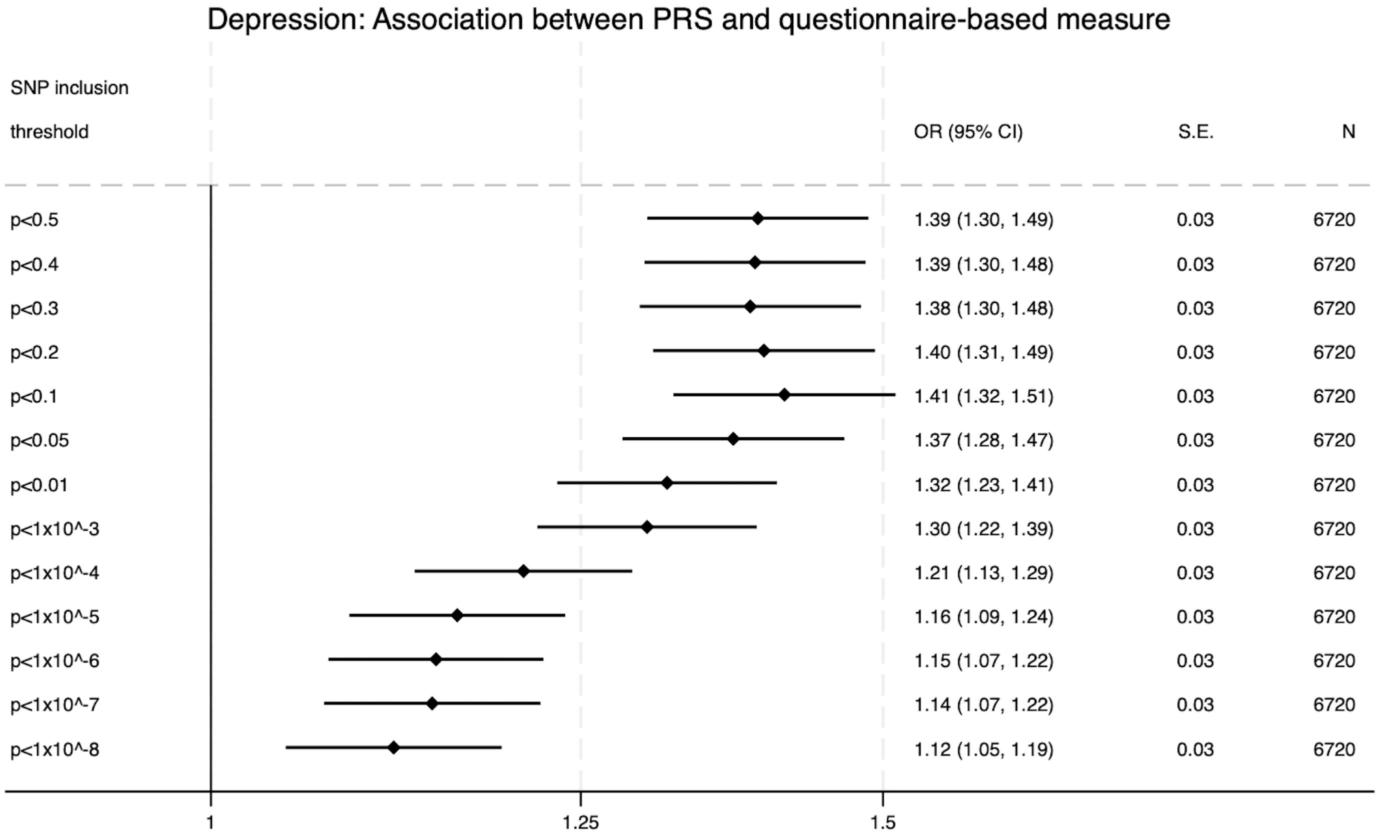

**Anxiety:**

Anxiety was measured at 18 weeks' gestation using the anxiety subscale of the Crown-Crisp Experimental Index (CCEI) [13], a validated self-rating inventory. A binary indicator was derived from the CCEI anxiety score, with a score of 9 or more indicating a high level of anxiety [14]. Thirteen logistic regression models were fitted, one for each SNP-inclusion p-value threshold. All ORs and most of the associated 95% CIs were above 1, indicating strong associations between anxiety PRSs and questionnaire-based anxiety.

#### Figure 3.2

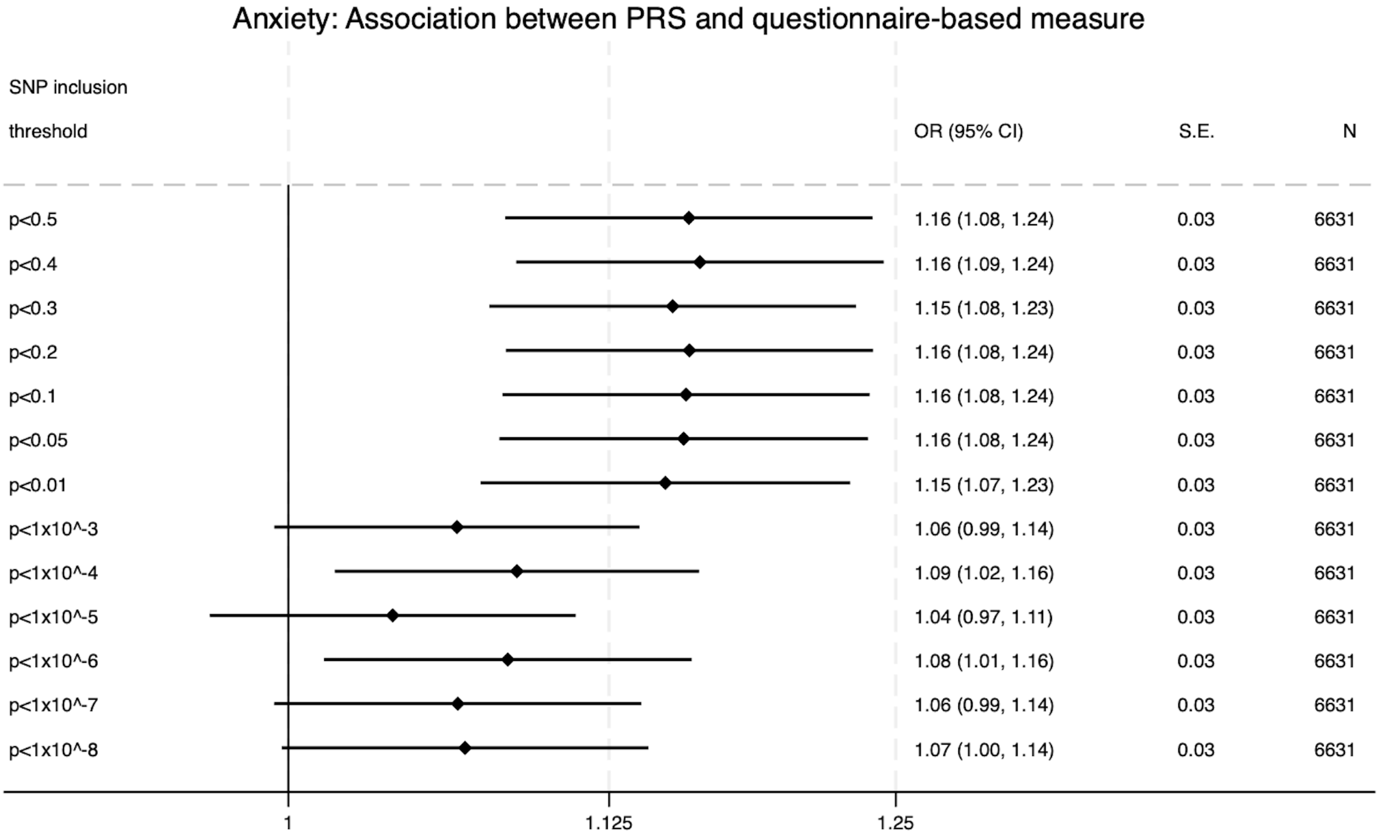

### 4. Primary analysis

Table 2 gives odds ratios (with 95% confidence intervals), unadjusted p-values, and FDR-adjusted p-values from the primary analysis.

#### Table 4.1

| Primary analysis | | | | | | | |
|---|---|---|---|---|---|---|---|
| **PRS** | **Urinary symptom** | **2002-04** | | | **2011-12** | | |
| | | OR (95% CI) | p | p (FDR) | OR (95% CI) | p | p (FDR) |
| Anxiety | Any UI | 1.08 (1.00-1.17) | 0.05 | 0.15 | 1.07 (0.99-1.16) | 0.10 | 0.19 |
| Anxiety | Any urgency | 1.08 (0.98-1.18) | 0.11 | 0.20 | 1.09 (0.99-1.19) | 0.08 | 0.19 |
| Anxiety | Mixed UI | 1.20 (1.03-1.39)* | 0.02 | 0.06 | 1.04 (0.91-1.18) | 0.59 | 0.64 |
| Anxiety | Nocturia | 1.12 (0.99-1.26) | 0.07 | 0.18 | 1.05 (0.93-1.18) | 0.46 | 0.54 |
| Anxiety | Stress UI | 1.01 (0.92-1.11) | 0.80 | 0.82 | 1.06 (0.96-1.18) | 0.24 | 0.34 |
| Anxiety | Urgency UI | 1.19 (0.94-1.51) | 0.16 | 0.26 | 1.12 (0.90-1.38) | 0.31 | 0.41 |
| Depression | Any UI | 1.09 (1.01-1.18)* | 0.03 | 0.09 | 1.11 (1.02-1.20)* | 0.02 | 0.06 |
| Depression | Any urgency | 1.08 (0.99-1.19) | 0.10 | 0.19 | 1.13 (1.03-1.24)** | 0.01 | 0.04 |
| Depression | Mixed UI | 1.10 (0.95-1.28) | 0.20 | 0.30 | 1.05 (0.93-1.20) | 0.43 | 0.54 |
| Depression | Nocturia | 1.33 (1.17-1.50)** | <0.01 | <0.01 | 1.24 (1.09-1.40)** | <0.01 | 0.01 |
| Depression | Stress UI | 1.09 (0.99-1.20) | 0.08 | 0.19 | 1.08 (0.97-1.19) | 0.16 | 0.26 |
| Depression | Urgency UI | 1.03 (0.81-1.31) | 0.82 | 0.82 | 1.24 (1.00-1.53) | 0.05 | 0.15 |
| Neuroticism | Any UI | 1.14 (1.05-1.23)** | <0.01 | 0.01 | 1.03 (0.95-1.12) | 0.45 | 0.54 |
| Neuroticism | Any urgency | 1.12 (1.02-1.23)* | 0.01 | 0.06 | 1.06 (0.97-1.17) | 0.19 | 0.29 |
| Neuroticism | Mixed UI | 1.23 (1.07-1.43)** | <0.01 | 0.03 | 0.98 (0.86-1.11) | 0.75 | 0.80 |
| Neuroticism | Nocturia | 1.24 (1.10-1.40)** | <0.01 | 0.01 | 1.21 (1.07-1.37)** | <0.01 | 0.02 |
| Neuroticism | Stress UI | 1.08 (0.98-1.18) | 0.12 | 0.21 | 1.05 (0.95-1.16) | 0.36 | 0.46 |
| Neuroticism | Urgency UI | 1.14 (0.90-1.44) | 0.27 | 0.37 | 1.07 (0.87-1.32) | 0.53 | 0.60 |

Key:
*Unadjusted p-value<0.05
**FDR-adjusted p-value<0.05

### 5. Sensitivity analysis

The sensitivity analysis investigated the same associations as the primary analysis but excluded subjects with organic causes of LUTS. Pelvic inflammatory disease, kidney disease, diabetes (non-gestational), and pregnancy were identified as organic causes of LUTS.

This data was available for 2002-04 so women with any of the above conditions were excluded for the sensitivity analysis. However, for the 2011-12 timepoint, the closest available data was collected in 2010. Data was collected on pregnancy and diabetes (not differentiating between gestational and non-gestational diabetes). Therefore, for the 2011-12 sensitivity analysis, women who were pregnant or diabetic as of 2010 were excluded.

190 subjects were excluded for the 2002-04 sensitivity analysis, whilst 103 subjects were excluded for the 2011-12 sensitivity analysis.

Table 3 and Table 4 give the breakdown of organic causes of LUTS for the subjects who were excluded at 2002-04 and 2011-12 respectively.

#### Table 5.1

| **Organic cause of LUTS*** | **Frequency (% out of N=190 excluded)** |
| --- | --- |
| Pelvic inflammatory disease | 23 (12.1%) |
| Diabetes (non-gestational) | 69 (36.3%) |
| Kidney disease | 57 (30.0%) |
| Pregnancy | 49 (25.8%) |

#### Table 5.2

| **Organic cause of LUTS*** | **Frequency (% out of N=103 excluded)** |
| --- | --- |
| Diabetes | 92 (89.3%) |
| Pregnancy | 12 (11.7%) |

*Note: Some subjects had more than one organic cause of LUTS

Figure 1 shows odds ratios, p-values and sample sizes for the analysis of PRS scores at the $p<0.05$ SNP-inclusion significance level, excluding subjects with organic causes of LUTS.

### Figure 5.1

#### Sensitivity analysis: Odds ratios (95% CI)

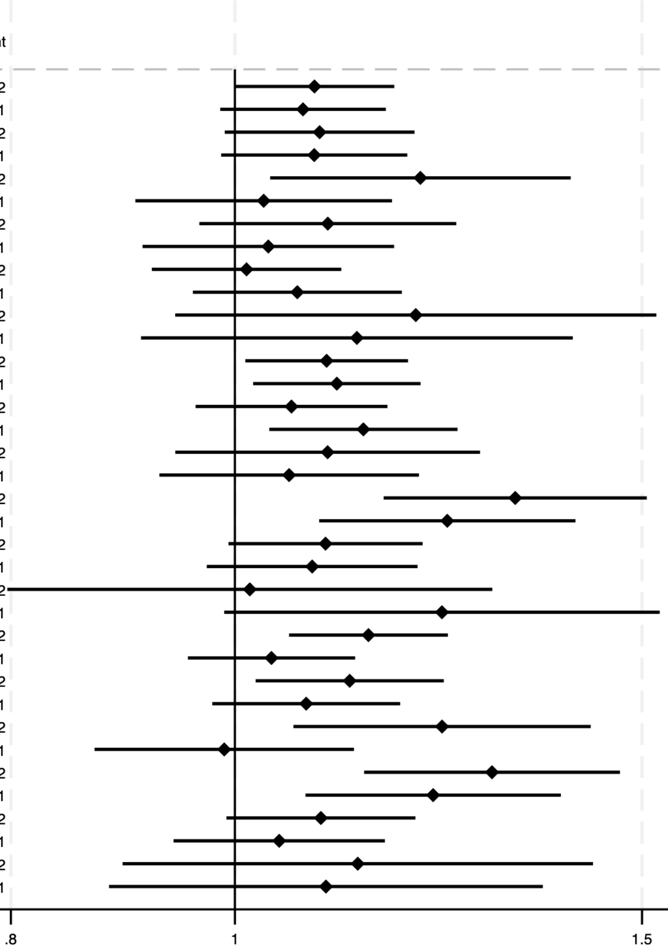

| PRS | Urinary symptom | Timepoint | OR (95% CI) | p | adj p | N | Case % |
|---|---|---|---|---|---|---|---|
| Anxiety | Any UI | 2002 | 1.08 (1.00, 1.17) | 0.05 | 0.15 | 4637 | 15.76 |
| Anxiety | Any UI | 2011 | 1.07 (0.99, 1.16) | 0.11 | 0.21 | 2968 | 24.76 |
| Anxiety | Any Urgency | 2002 | 1.09 (0.99, 1.20) | 0.08 | 0.18 | 4637 | 10.44 |
| Anxiety | Any Urgency | 2011 | 1.08 (0.99, 1.19) | 0.10 | 0.20 | 2968 | 17.96 |
| Anxiety | Mixed UI | 2002 | 1.20 (1.04, 1.40) | 0.02 | 0.06 | 4637 | 3.90 |
| Anxiety | Mixed UI | 2011 | 1.03 (0.91, 1.17) | 0.66 | 0.72 | 2968 | 8.46 |
| Anxiety | Nocturia | 2002 | 1.10 (0.97, 1.25) | 0.16 | 0.26 | 4637 | 5.37 |
| Anxiety | Nocturia | 2011 | 1.03 (0.91, 1.17) | 0.60 | 0.68 | 2968 | 8.86 |
| Anxiety | Stress UI | 2002 | 1.01 (0.92, 1.11) | 0.81 | 0.86 | 4637 | 10.37 |
| Anxiety | Stress UI | 2011 | 1.06 (0.96, 1.18) | 0.24 | 0.36 | 2968 | 13.48 |
| Anxiety | Urgency UI | 2002 | 1.20 (0.94, 1.52) | 0.14 | 0.25 | 4637 | 1.49 |
| Anxiety | Urgency UI | 2011 | 1.13 (0.91, 1.40) | 0.27 | 0.37 | 2968 | 2.83 |
| Depression | Any UI | 2002 | 1.10 (1.01, 1.19) | 0.03 | 0.09 | 4637 | 15.76 |
| Depression | Any UI | 2011 | 1.11 (1.02, 1.20) | 0.02 | 0.06 | 2968 | 24.76 |
| Depression | Any Urgency | 2002 | 1.06 (0.96, 1.16) | 0.25 | 0.36 | 4637 | 10.44 |
| Depression | Any Urgency | 2011 | 1.14 (1.03, 1.25) | 0.01 | 0.04 | 2968 | 17.96 |
| Depression | Mixed UI | 2002 | 1.10 (0.94, 1.28) | 0.23 | 0.36 | 4637 | 3.90 |
| Depression | Mixed UI | 2011 | 1.06 (0.93, 1.20) | 0.41 | 0.48 | 2968 | 8.46 |
| Depression | Nocturia | 2002 | 1.32 (1.16, 1.51) | 0.00 | 0.00 | 4637 | 5.37 |
| Depression | Nocturia | 2011 | 1.24 (1.09, 1.40) | 0.00 | 0.01 | 2968 | 8.86 |
| Depression | Stress UI | 2002 | 1.09 (0.99, 1.21) | 0.07 | 0.17 | 4637 | 10.37 |
| Depression | Stress UI | 2011 | 1.08 (0.97, 1.20) | 0.15 | 0.26 | 2968 | 13.48 |
| Depression | Urgency UI | 2002 | 1.01 (0.80, 1.29) | 0.90 | 0.90 | 4637 | 1.49 |
| Depression | Urgency UI | 2011 | 1.23 (0.99, 1.53) | 0.06 | 0.17 | 2968 | 2.83 |
| Neuroticism | Any UI | 2002 | 1.14 (1.06, 1.24) | 0.00 | 0.01 | 4637 | 15.76 |
| Neuroticism | Any UI | 2011 | 1.04 (0.95, 1.13) | 0.39 | 0.48 | 2968 | 24.76 |
| Neuroticism | Any Urgency | 2002 | 1.12 (1.02, 1.23) | 0.02 | 0.06 | 4637 | 10.44 |
| Neuroticism | Any Urgency | 2011 | 1.07 (0.98, 1.18) | 0.14 | 0.25 | 2968 | 17.96 |
| Neuroticism | Mixed UI | 2002 | 1.23 (1.06, 1.43) | 0.01 | 0.04 | 4637 | 3.90 |
| Neuroticism | Mixed UI | 2011 | 0.99 (0.87, 1.13) | 0.87 | 0.90 | 2968 | 8.46 |
| Neuroticism | Nocturia | 2002 | 1.29 (1.14, 1.47) | 0.00 | 0.00 | 4637 | 5.37 |
| Neuroticism | Nocturia | 2011 | 1.22 (1.07, 1.38) | 0.00 | 0.02 | 2968 | 8.86 |
| Neuroticism | Stress UI | 2002 | 1.09 (0.99, 1.20) | 0.07 | 0.18 | 4637 | 10.37 |
| Neuroticism | Stress UI | 2011 | 1.05 (0.94, 1.16) | 0.41 | 0.48 | 2968 | 13.48 |
| Neuroticism | Urgency UI | 2002 | 1.13 (0.89, 1.43) | 0.31 | 0.41 | 4637 | 1.49 |
| Neuroticism | Urgency UI | 2011 | 1.09 (0.88, 1.36) | 0.41 | 0.48 | 2968 | 2.83 |

### 6. PRS validation

Figures 2-13 show associations between LUTS and PRSs at each of the SNP-inclusion significance thresholds ranging from $p<0.5$ to $p<5x10^{-8}$ (S1 to S13). Results for the $p<0.05$ threshold (S6) are omitted as these form the primary results which are included in the main paper. Table 5 gives the key matching each analysis to its respective significance threshold.

Each figure reports odds ratios with 95% confidence intervals and unadjusted p-values. The figures also give the number of subjects in each model as well as the percentage of subjects who were positive cases for the urinary symptom.

#### Table 6.1

| Analysis | SNP-inclusion significance threshold |
|---|---|
| S1 | $p<0.5$ |
| S2 | $p<0.4$ |
| S3 | $p<0.3$ |
| S4 | $p<0.2$ |
| S5 | $p<0.1$ |
| S6* | $p<0.05$ |
| S7 | $p<0.01$ |
| S8 | $p<1x10^{-3}$ |
| S9 | $p<1x10^{-4}$ |
| S10 | $p<1x10^{-5}$ |
| S11 | $p<1x10^{-6}$ |
| S12 | $p<1x10^{-7}$ |
| S13 | $p<5x10^{-8}$ |

*S6 analysis omitted from the supplement as this is the primary result which can be found in the main paper.

#### Figure 6.1

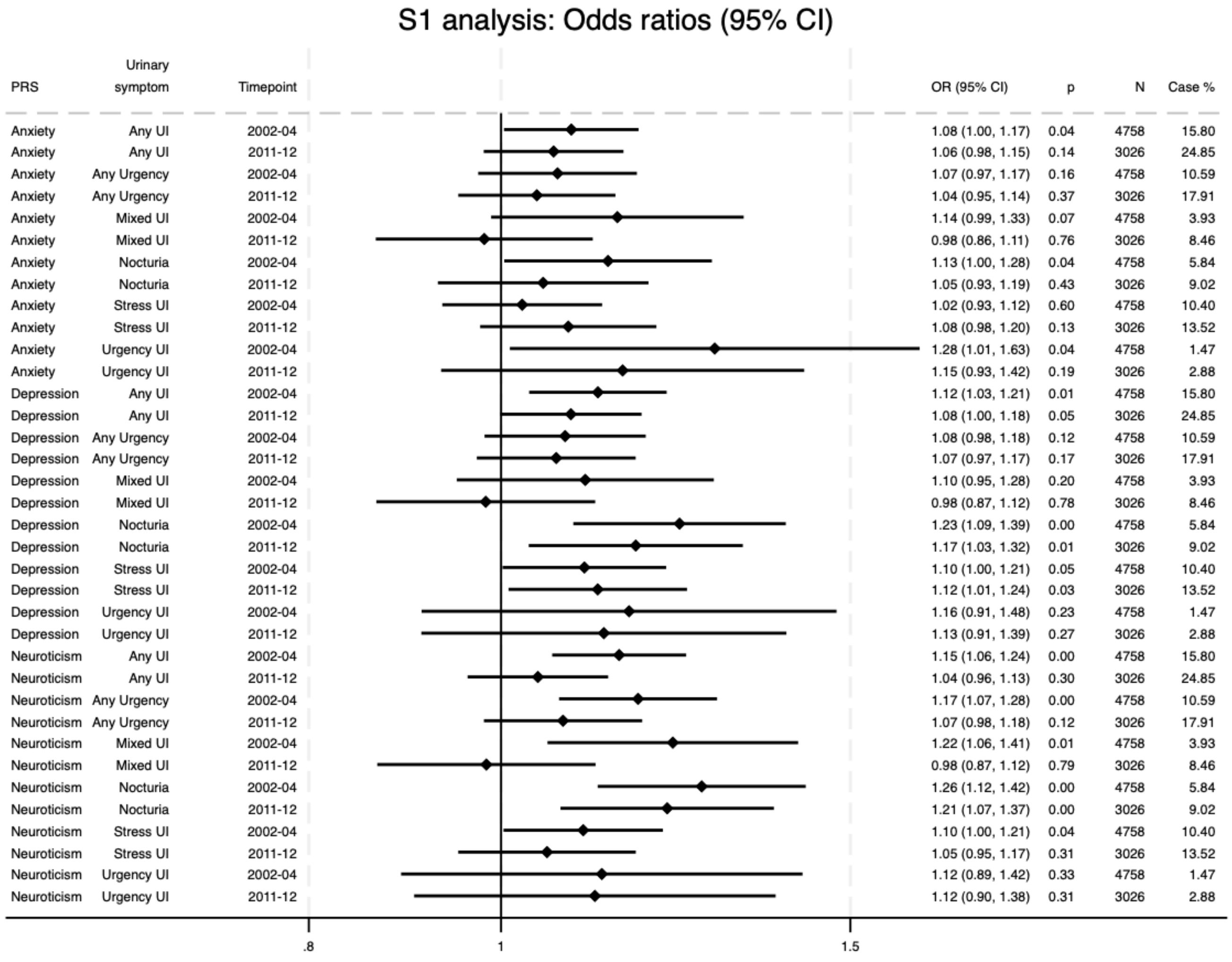
S1 analysis: Odds ratios (95% CI)
PRS
Urinary symptom
Timepoint
OR (95% CI)
p
N
Case %
Anxiety Any UI 2002-04 1.08 (1.00, 1.17) 0.04 4758 15.80
Anxiety Any UI 2011-12 1.06 (0.98, 1.15) 0.14 3026 24.85
Anxiety Any Urgency 2002-04 1.07 (0.97, 1.17) 0.16 4758 10.59
Anxiety Any Urgency 2011-12 1.04 (0.95, 1.14) 0.37 3026 17.91
Anxiety Mixed UI 2002-04 1.14 (0.99, 1.33) 0.07 4758 3.93
Anxiety Mixed UI 2011-12 0.98 (0.86, 1.11) 0.76 3026 8.46
Anxiety Nocturia 2002-04 1.13 (1.00, 1.28) 0.04 4758 5.84
Anxiety Nocturia 2011-12 1.05 (0.93, 1.19) 0.43 3026 9.02
Anxiety Stress UI 2002-04 1.02 (0.93, 1.12) 0.60 4758 10.40
Anxiety Stress UI 2011-12 1.08 (0.98, 1.20) 0.13 3026 13.52
Anxiety Urgency UI 2002-04 1.28 (1.01, 1.63) 0.04 4758 1.47
Anxiety Urgency UI 2011-12 1.15 (0.93, 1.42) 0.19 3026 2.88
Depression Any UI 2002-04 1.12 (1.03, 1.21) 0.01 4758 15.80
Depression Any UI 2011-12 1.08 (1.00, 1.18) 0.05 3026 24.85
Depression Any Urgency 2002-04 1.08 (0.98, 1.18) 0.12 4758 10.59
Depression Any Urgency 2011-12 1.07 (0.97, 1.17) 0.17 3026 17.91
Depression Mixed UI 2002-04 1.10 (0.95, 1.28) 0.20 4758 3.93
Depression Mixed UI 2011-12 0.98 (0.87, 1.12) 0.78 3026 8.46
Depression Nocturia 2002-04 1.23 (1.09, 1.39) 0.00 4758 5.84
Depression Nocturia 2011-12 1.17 (1.03, 1.32) 0.01 3026 9.02
Depression Stress UI 2002-04 1.10 (1.00, 1.21) 0.05 4758 10.40
Depression Stress UI 2011-12 1.12 (1.01, 1.24) 0.03 3026 13.52
Depression Urgency UI 2002-04 1.16 (0.91, 1.48) 0.23 4758 1.47
Depression Urgency UI 2011-12 1.13 (0.91, 1.39) 0.27 3026 2.88
Neuroticism Any UI 2002-04 1.15 (1.06, 1.24) 0.00 4758 15.80
Neuroticism Any UI 2011-12 1.04 (0.96, 1.13) 0.30 3026 24.85
Neuroticism Any Urgency 2002-04 1.17 (1.07, 1.28) 0.00 4758 10.59
Neuroticism Any Urgency 2011-12 1.07 (0.98, 1.18) 0.12 3026 17.91
Neuroticism Mixed UI 2002-04 1.22 (1.06, 1.41) 0.01 4758 3.93
Neuroticism Mixed UI 2011-12 0.98 (0.87, 1.12) 0.79 3026 8.46
Neuroticism Nocturia 2002-04 1.26 (1.12, 1.42) 0.00 4758 5.84
Neuroticism Nocturia 2011-12 1.21 (1.07, 1.37) 0.00 3026 9.02
Neuroticism Stress UI 2002-04 1.10 (1.00, 1.21) 0.04 4758 10.40
Neuroticism Stress UI 2011-12 1.05 (0.95, 1.17) 0.31 3026 13.52
Neuroticism Urgency UI 2002-04 1.12 (0.89, 1.42) 0.33 4758 1.47
Neuroticism Urgency UI 2011-12 1.12 (0.90, 1.38) 0.31 3026 2.88
.8
1
1.5

#### Figure 6.2

S2 analysis: Odds ratios (95% CI)

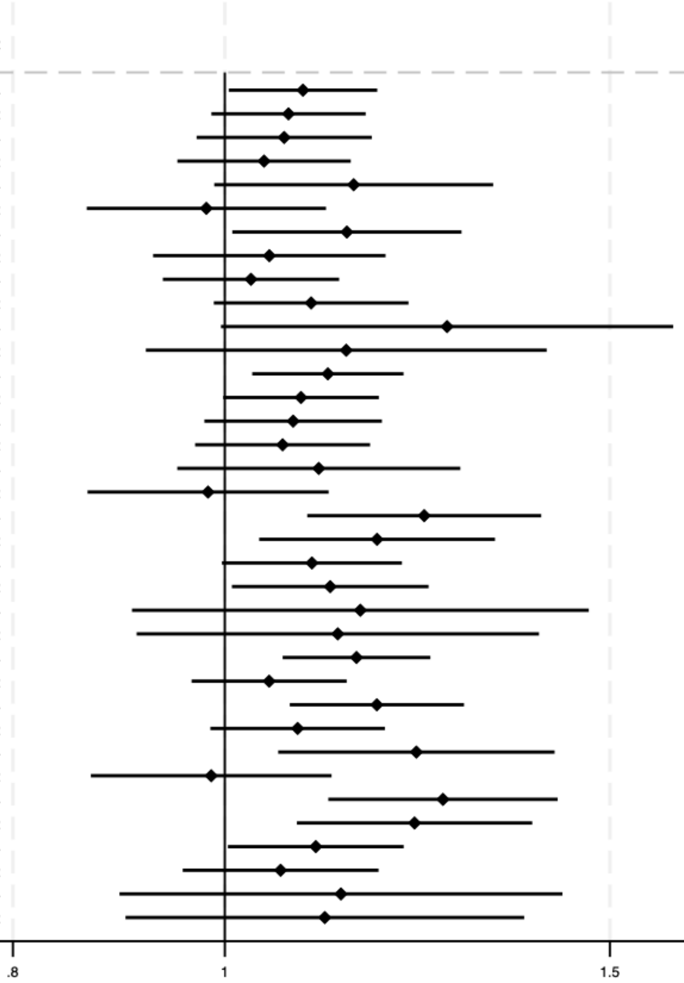

| PRS | Urinary symptom | Timepoint | OR (95% CI) | p | N | Case % |
|---|---|---|---|---|---|---|
| Anxiety | Any UI | 2002-04 | 1.09 (1.00, 1.17) | 0.04 | 4758 | 15.80 |
| Anxiety | Any UI | 2011-12 | 1.07 (0.99, 1.16) | 0.11 | 3026 | 24.85 |
| Anxiety | Any Urgency | 2002-04 | 1.06 (0.97, 1.17) | 0.18 | 4758 | 10.59 |
| Anxiety | Any Urgency | 2011-12 | 1.04 (0.95, 1.14) | 0.38 | 3026 | 17.91 |
| Anxiety | Mixed UI | 2002-04 | 1.15 (0.99, 1.33) | 0.07 | 4758 | 3.93 |
| Anxiety | Mixed UI | 2011-12 | 0.98 (0.86, 1.11) | 0.76 | 3026 | 8.46 |
| Anxiety | Nocturia | 2002-04 | 1.14 (1.01, 1.28) | 0.04 | 4758 | 5.84 |
| Anxiety | Nocturia | 2011-12 | 1.05 (0.93, 1.18) | 0.45 | 3026 | 9.02 |
| Anxiety | Stress UI | 2002-04 | 1.03 (0.94, 1.13) | 0.56 | 4758 | 10.40 |
| Anxiety | Stress UI | 2011-12 | 1.10 (0.99, 1.21) | 0.08 | 3026 | 13.52 |
| Anxiety | Urgency UI | 2002-04 | 1.26 (1.00, 1.60) | 0.05 | 4758 | 1.47 |
| Anxiety | Urgency UI | 2011-12 | 1.14 (0.92, 1.40) | 0.23 | 3026 | 2.88 |
| Depression | Any UI | 2002-04 | 1.11 (1.03, 1.21) | 0.01 | 4758 | 15.80 |
| Depression | Any UI | 2011-12 | 1.08 (1.00, 1.18) | 0.06 | 3026 | 24.85 |
| Depression | Any Urgency | 2002-04 | 1.07 (0.98, 1.18) | 0.13 | 4758 | 10.59 |
| Depression | Any Urgency | 2011-12 | 1.06 (0.97, 1.17) | 0.20 | 3026 | 17.91 |
| Depression | Mixed UI | 2002-04 | 1.10 (0.95, 1.28) | 0.19 | 4758 | 3.93 |
| Depression | Mixed UI | 2011-12 | 0.98 (0.87, 1.12) | 0.79 | 3026 | 8.46 |
| Depression | Nocturia | 2002-04 | 1.23 (1.09, 1.40) | 0.00 | 4758 | 5.84 |
| Depression | Nocturia | 2011-12 | 1.17 (1.04, 1.33) | 0.01 | 3026 | 9.02 |
| Depression | Stress UI | 2002-04 | 1.10 (1.00, 1.21) | 0.06 | 4758 | 10.40 |
| Depression | Stress UI | 2011-12 | 1.12 (1.01, 1.24) | 0.04 | 3026 | 13.52 |
| Depression | Urgency UI | 2002-04 | 1.15 (0.91, 1.47) | 0.24 | 4758 | 1.47 |
| Depression | Urgency UI | 2011-12 | 1.13 (0.91, 1.39) | 0.27 | 3026 | 2.88 |
| Neuroticism | Any UI | 2002-04 | 1.15 (1.06, 1.24) | 0.00 | 4758 | 15.80 |
| Neuroticism | Any UI | 2011-12 | 1.05 (0.97, 1.14) | 0.26 | 3026 | 24.85 |
| Neuroticism | Any Urgency | 2002-04 | 1.17 (1.07, 1.29) | 0.00 | 4758 | 10.59 |
| Neuroticism | Any Urgency | 2011-12 | 1.08 (0.98, 1.18) | 0.10 | 3026 | 17.91 |
| Neuroticism | Mixed UI | 2002-04 | 1.22 (1.06, 1.42) | 0.01 | 4758 | 3.93 |
| Neuroticism | Mixed UI | 2011-12 | 0.99 (0.87, 1.12) | 0.82 | 3026 | 8.46 |
| Neuroticism | Nocturia | 2002-04 | 1.26 (1.12, 1.42) | 0.00 | 4758 | 5.84 |
| Neuroticism | Nocturia | 2011-12 | 1.22 (1.08, 1.38) | 0.00 | 3026 | 9.02 |
| Neuroticism | Stress UI | 2002-04 | 1.10 (1.00, 1.21) | 0.04 | 4758 | 10.40 |
| Neuroticism | Stress UI | 2011-12 | 1.06 (0.96, 1.18) | 0.26 | 3026 | 13.52 |
| Neuroticism | Urgency UI | 2002-04 | 1.13 (0.89, 1.43) | 0.30 | 4758 | 1.47 |
| Neuroticism | Urgency UI | 2011-12 | 1.11 (0.90, 1.37) | 0.33 | 3026 | 2.88 |

### Figure 6.3

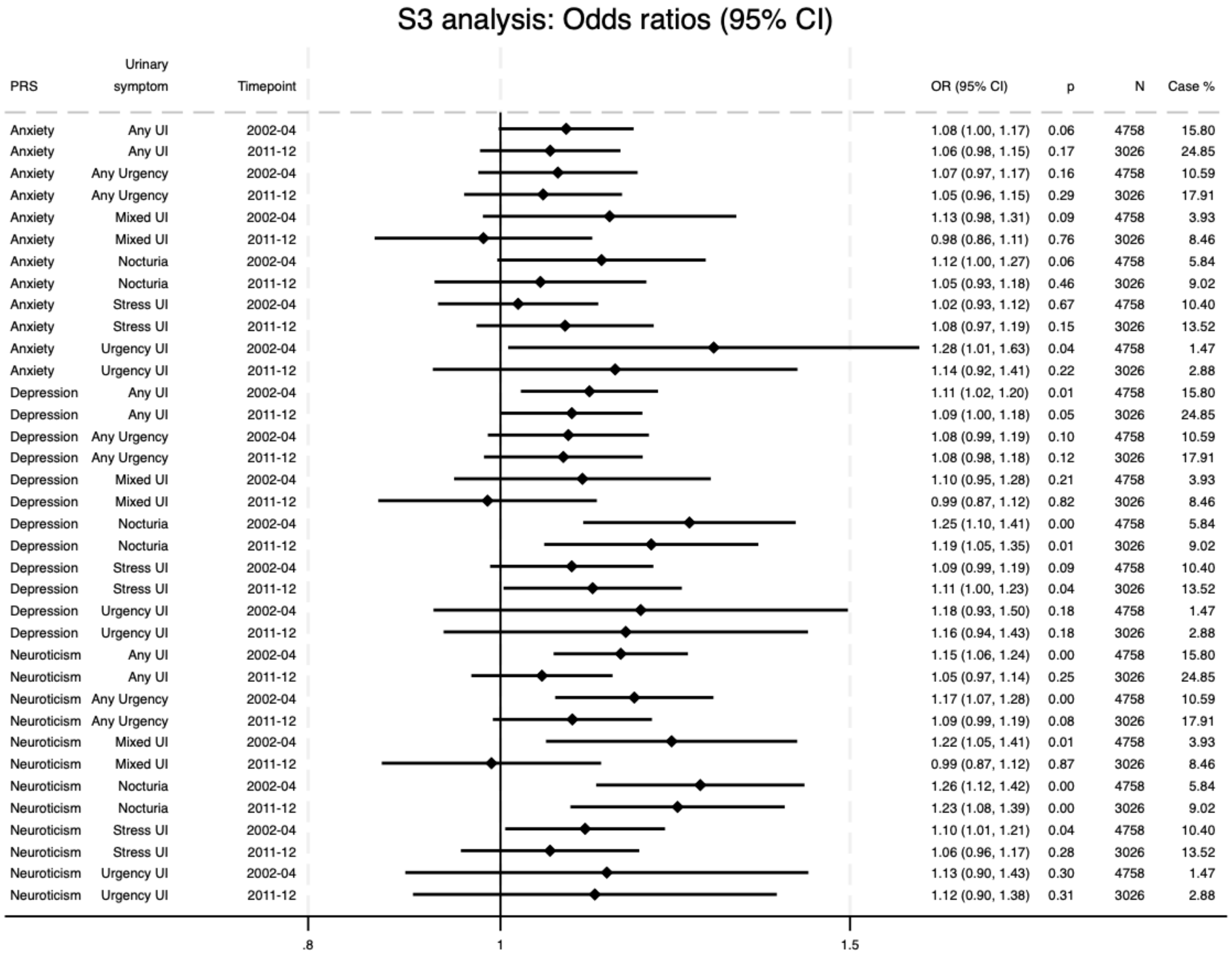
S3 analysis: Odds ratios (95% CI)
PRS | Urinary symptom | Timepoint | OR (95% CI) | p | N | Case %
Anxiety | Any UI | 2002-04 | 1.08 (1.00, 1.17) | 0.06 | 4758 | 15.80
Anxiety | Any UI | 2011-12 | 1.06 (0.98, 1.15) | 0.17 | 3026 | 24.85
Anxiety | Any Urgency | 2002-04 | 1.07 (0.97, 1.17) | 0.16 | 4758 | 10.59
Anxiety | Any Urgency | 2011-12 | 1.05 (0.96, 1.15) | 0.29 | 3026 | 17.91
Anxiety | Mixed UI | 2002-04 | 1.13 (0.98, 1.31) | 0.09 | 4758 | 3.93
Anxiety | Mixed UI | 2011-12 | 0.98 (0.86, 1.11) | 0.76 | 3026 | 8.46
Anxiety | Nocturia | 2002-04 | 1.12 (1.00, 1.27) | 0.06 | 4758 | 5.84
Anxiety | Nocturia | 2011-12 | 1.05 (0.93, 1.18) | 0.46 | 3026 | 9.02
Anxiety | Stress UI | 2002-04 | 1.02 (0.93, 1.12) | 0.67 | 4758 | 10.40
Anxiety | Stress UI | 2011-12 | 1.08 (0.97, 1.19) | 0.15 | 3026 | 13.52
Anxiety | Urgency UI | 2002-04 | 1.28 (1.01, 1.63) | 0.04 | 4758 | 1.47
Anxiety | Urgency UI | 2011-12 | 1.14 (0.92, 1.41) | 0.22 | 3026 | 2.88
Depression | Any UI | 2002-04 | 1.11 (1.02, 1.20) | 0.01 | 4758 | 15.80
Depression | Any UI | 2011-12 | 1.09 (1.00, 1.18) | 0.05 | 3026 | 24.85
Depression | Any Urgency | 2002-04 | 1.08 (0.99, 1.19) | 0.10 | 4758 | 10.59
Depression | Any Urgency | 2011-12 | 1.08 (0.98, 1.18) | 0.12 | 3026 | 17.91
Depression | Mixed UI | 2002-04 | 1.10 (0.95, 1.28) | 0.21 | 4758 | 3.93
Depression | Mixed UI | 2011-12 | 0.99 (0.87, 1.12) | 0.82 | 3026 | 8.46
Depression | Nocturia | 2002-04 | 1.25 (1.10, 1.41) | 0.00 | 4758 | 5.84
Depression | Nocturia | 2011-12 | 1.19 (1.05, 1.35) | 0.01 | 3026 | 9.02
Depression | Stress UI | 2002-04 | 1.09 (0.99, 1.19) | 0.09 | 4758 | 10.40
Depression | Stress UI | 2011-12 | 1.11 (1.00, 1.23) | 0.04 | 3026 | 13.52
Depression | Urgency UI | 2002-04 | 1.18 (0.93, 1.50) | 0.18 | 4758 | 1.47
Depression | Urgency UI | 2011-12 | 1.16 (0.94, 1.43) | 0.18 | 3026 | 2.88
Neuroticism | Any UI | 2002-04 | 1.15 (1.06, 1.24) | 0.00 | 4758 | 15.80
Neuroticism | Any UI | 2011-12 | 1.05 (0.97, 1.14) | 0.25 | 3026 | 24.85
Neuroticism | Any Urgency | 2002-04 | 1.17 (1.07, 1.28) | 0.00 | 4758 | 10.59
Neuroticism | Any Urgency | 2011-12 | 1.09 (0.99, 1.19) | 0.08 | 3026 | 17.91
Neuroticism | Mixed UI | 2002-04 | 1.22 (1.05, 1.41) | 0.01 | 4758 | 3.93
Neuroticism | Mixed UI | 2011-12 | 0.99 (0.87, 1.12) | 0.87 | 3026 | 8.46
Neuroticism | Nocturia | 2002-04 | 1.26 (1.12, 1.42) | 0.00 | 4758 | 5.84
Neuroticism | Nocturia | 2011-12 | 1.23 (1.08, 1.39) | 0.00 | 3026 | 9.02
Neuroticism | Stress UI | 2002-04 | 1.10 (1.01, 1.21) | 0.04 | 4758 | 10.40
Neuroticism | Stress UI | 2011-12 | 1.06 (0.96, 1.17) | 0.28 | 3026 | 13.52
Neuroticism | Urgency UI | 2002-04 | 1.13 (0.90, 1.43) | 0.30 | 4758 | 1.47
Neuroticism | Urgency UI | 2011-12 | 1.12 (0.90, 1.38) | 0.31 | 3026 | 2.88
.8
1
1.5

### Figure 6.4

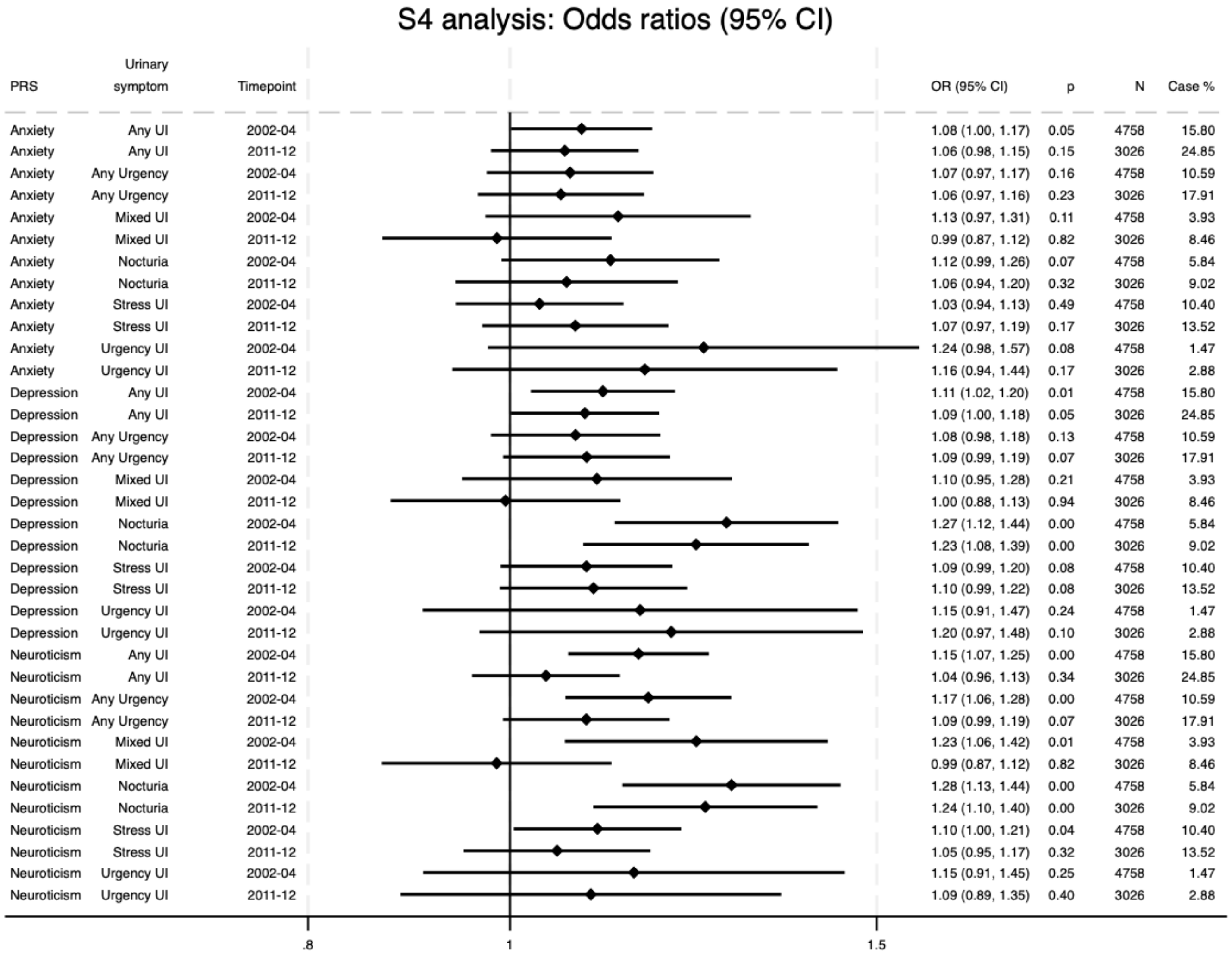
S4 analysis: Odds ratios (95% CI)
PRS
Urinary symptom
Timepoint
OR (95% CI)
p
N
Case %
Anxiety Any UI 2002-04 1.08 (1.00, 1.17) 0.05 4758 15.80
Anxiety Any UI 2011-12 1.06 (0.98, 1.15) 0.15 3026 24.85
Anxiety Any Urgency 2002-04 1.07 (0.97, 1.17) 0.16 4758 10.59
Anxiety Any Urgency 2011-12 1.06 (0.97, 1.16) 0.23 3026 17.91
Anxiety Mixed UI 2002-04 1.13 (0.97, 1.31) 0.11 4758 3.93
Anxiety Mixed UI 2011-12 0.99 (0.87, 1.12) 0.82 3026 8.46
Anxiety Nocturia 2002-04 1.12 (0.99, 1.26) 0.07 4758 5.84
Anxiety Nocturia 2011-12 1.06 (0.94, 1.20) 0.32 3026 9.02
Anxiety Stress UI 2002-04 1.03 (0.94, 1.13) 0.49 4758 10.40
Anxiety Stress UI 2011-12 1.07 (0.97, 1.19) 0.17 3026 13.52
Anxiety Urgency UI 2002-04 1.24 (0.98, 1.57) 0.08 4758 1.47
Anxiety Urgency UI 2011-12 1.16 (0.94, 1.44) 0.17 3026 2.88
Depression Any UI 2002-04 1.11 (1.02, 1.20) 0.01 4758 15.80
Depression Any UI 2011-12 1.09 (1.00, 1.18) 0.05 3026 24.85
Depression Any Urgency 2002-04 1.08 (0.98, 1.18) 0.13 4758 10.59
Depression Any Urgency 2011-12 1.09 (0.99, 1.19) 0.07 3026 17.91
Depression Mixed UI 2002-04 1.10 (0.95, 1.28) 0.21 4758 3.93
Depression Mixed UI 2011-12 1.00 (0.88, 1.13) 0.94 3026 8.46
Depression Nocturia 2002-04 1.27 (1.12, 1.44) 0.00 4758 5.84
Depression Nocturia 2011-12 1.23 (1.08, 1.39) 0.00 3026 9.02
Depression Stress UI 2002-04 1.09 (0.99, 1.20) 0.08 4758 10.40
Depression Stress UI 2011-12 1.10 (0.99, 1.22) 0.08 3026 13.52
Depression Urgency UI 2002-04 1.15 (0.91, 1.47) 0.24 4758 1.47
Depression Urgency UI 2011-12 1.20 (0.97, 1.48) 0.10 3026 2.88
Neuroticism Any UI 2002-04 1.15 (1.07, 1.25) 0.00 4758 15.80
Neuroticism Any UI 2011-12 1.04 (0.96, 1.13) 0.34 3026 24.85
Neuroticism Any Urgency 2002-04 1.17 (1.06, 1.28) 0.00 4758 10.59
Neuroticism Any Urgency 2011-12 1.09 (0.99, 1.19) 0.07 3026 17.91
Neuroticism Mixed UI 2002-04 1.23 (1.06, 1.42) 0.01 4758 3.93
Neuroticism Mixed UI 2011-12 0.99 (0.87, 1.12) 0.82 3026 8.46
Neuroticism Nocturia 2002-04 1.28 (1.13, 1.44) 0.00 4758 5.84
Neuroticism Nocturia 2011-12 1.24 (1.10, 1.40) 0.00 3026 9.02
Neuroticism Stress UI 2002-04 1.10 (1.00, 1.21) 0.04 4758 10.40
Neuroticism Stress UI 2011-12 1.05 (0.95, 1.17) 0.32 3026 13.52
Neuroticism Urgency UI 2002-04 1.15 (0.91, 1.45) 0.25 4758 1.47
Neuroticism Urgency UI 2011-12 1.09 (0.89, 1.35) 0.40 3026 2.88
.8
1
1.5

### Figure 6.5

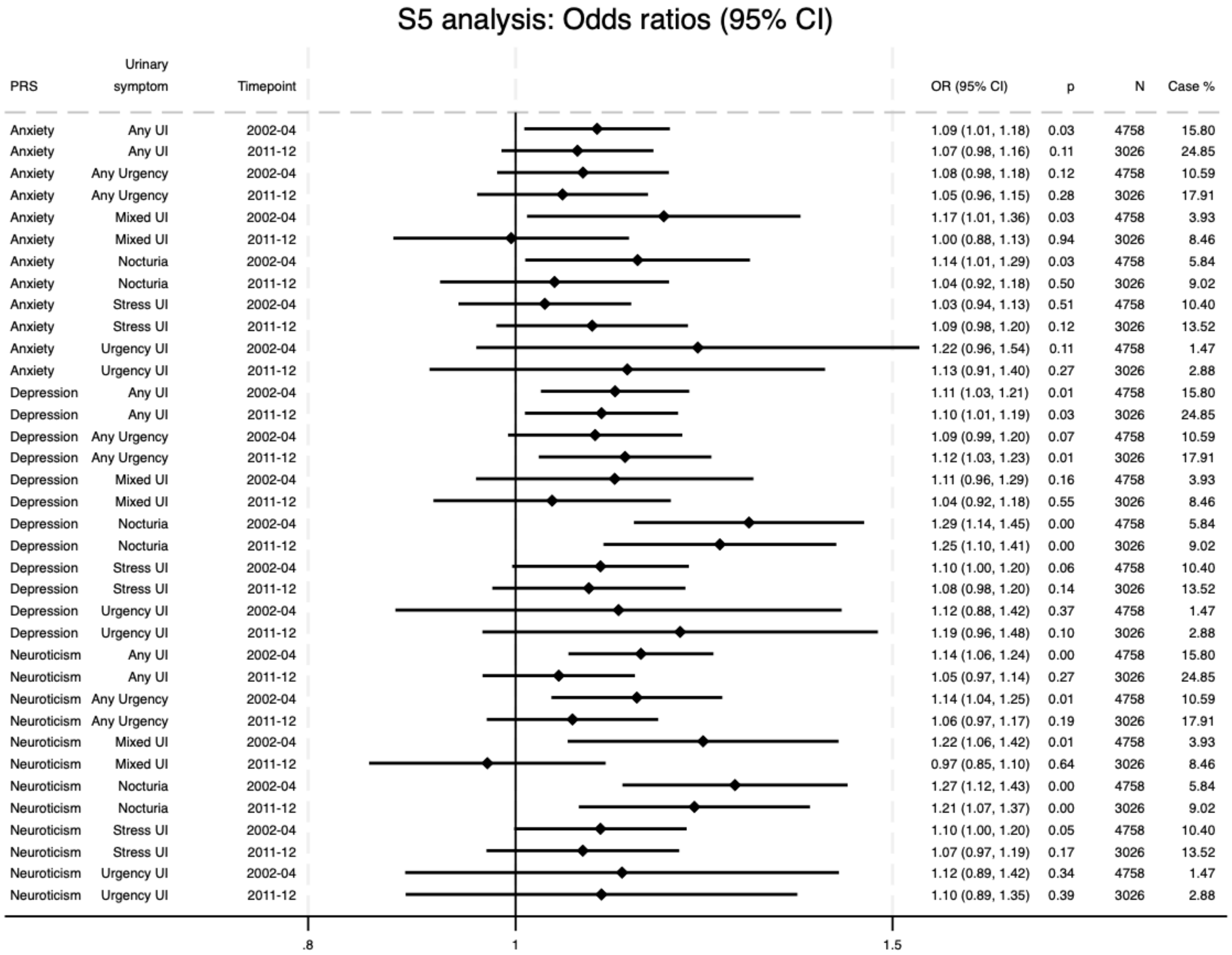
S5 analysis: Odds ratios (95% CI)
PRS | Urinary symptom | Timepoint | OR (95% CI) | p | N | Case %
Anxiety | Any UI | 2002-04 | 1.09 (1.01, 1.18) | 0.03 | 4758 | 15.80
Anxiety | Any UI | 2011-12 | 1.07 (0.98, 1.16) | 0.11 | 3026 | 24.85
Anxiety | Any Urgency | 2002-04 | 1.08 (0.98, 1.18) | 0.12 | 4758 | 10.59
Anxiety | Any Urgency | 2011-12 | 1.05 (0.96, 1.15) | 0.28 | 3026 | 17.91
Anxiety | Mixed UI | 2002-04 | 1.17 (1.01, 1.36) | 0.03 | 4758 | 3.93
Anxiety | Mixed UI | 2011-12 | 1.00 (0.88, 1.13) | 0.94 | 3026 | 8.46
Anxiety | Nocturia | 2002-04 | 1.14 (1.01, 1.29) | 0.03 | 4758 | 5.84
Anxiety | Nocturia | 2011-12 | 1.04 (0.92, 1.18) | 0.50 | 3026 | 9.02
Anxiety | Stress UI | 2002-04 | 1.03 (0.94, 1.13) | 0.51 | 4758 | 10.40
Anxiety | Stress UI | 2011-12 | 1.09 (0.98, 1.20) | 0.12 | 3026 | 13.52
Anxiety | Urgency UI | 2002-04 | 1.22 (0.96, 1.54) | 0.11 | 4758 | 1.47
Anxiety | Urgency UI | 2011-12 | 1.13 (0.91, 1.40) | 0.27 | 3026 | 2.88
Depression | Any UI | 2002-04 | 1.11 (1.03, 1.21) | 0.01 | 4758 | 15.80
Depression | Any UI | 2011-12 | 1.10 (1.01, 1.19) | 0.03 | 3026 | 24.85
Depression | Any Urgency | 2002-04 | 1.09 (0.99, 1.20) | 0.07 | 4758 | 10.59
Depression | Any Urgency | 2011-12 | 1.12 (1.03, 1.23) | 0.01 | 3026 | 17.91
Depression | Mixed UI | 2002-04 | 1.11 (0.96, 1.29) | 0.16 | 4758 | 3.93
Depression | Mixed UI | 2011-12 | 1.04 (0.92, 1.18) | 0.55 | 3026 | 8.46
Depression | Nocturia | 2002-04 | 1.29 (1.14, 1.45) | 0.00 | 4758 | 5.84
Depression | Nocturia | 2011-12 | 1.25 (1.10, 1.41) | 0.00 | 3026 | 9.02
Depression | Stress UI | 2002-04 | 1.10 (1.00, 1.20) | 0.06 | 4758 | 10.40
Depression | Stress UI | 2011-12 | 1.08 (0.98, 1.20) | 0.14 | 3026 | 13.52
Depression | Urgency UI | 2002-04 | 1.12 (0.88, 1.42) | 0.37 | 4758 | 1.47
Depression | Urgency UI | 2011-12 | 1.19 (0.96, 1.48) | 0.10 | 3026 | 2.88
Neuroticism | Any UI | 2002-04 | 1.14 (1.06, 1.24) | 0.00 | 4758 | 15.80
Neuroticism | Any UI | 2011-12 | 1.05 (0.97, 1.14) | 0.27 | 3026 | 24.85
Neuroticism | Any Urgency | 2002-04 | 1.14 (1.04, 1.25) | 0.01 | 4758 | 10.59
Neuroticism | Any Urgency | 2011-12 | 1.06 (0.97, 1.17) | 0.19 | 3026 | 17.91
Neuroticism | Mixed UI | 2002-04 | 1.22 (1.06, 1.42) | 0.01 | 4758 | 3.93
Neuroticism | Mixed UI | 2011-12 | 0.97 (0.85, 1.10) | 0.64 | 3026 | 8.46
Neuroticism | Nocturia | 2002-04 | 1.27 (1.12, 1.43) | 0.00 | 4758 | 5.84
Neuroticism | Nocturia | 2011-12 | 1.21 (1.07, 1.37) | 0.00 | 3026 | 9.02
Neuroticism | Stress UI | 2002-04 | 1.10 (1.00, 1.20) | 0.05 | 4758 | 10.40
Neuroticism | Stress UI | 2011-12 | 1.07 (0.97, 1.19) | 0.17 | 3026 | 13.52
Neuroticism | Urgency UI | 2002-04 | 1.12 (0.89, 1.42) | 0.34 | 4758 | 1.47
Neuroticism | Urgency UI | 2011-12 | 1.10 (0.89, 1.35) | 0.39 | 3026 | 2.88
.8
1
1.5

### Figure 6.6

#### S7 analysis: Odds ratios (95% CI)

| PRS | Urinary symptom | Timepoint | OR (95% CI) | p | N | Case % |
|---|---|---|---|---|---|---|
| Anxiety | Any UI | 2002-04 | 1.11 (1.03, 1.21) | 0.01 | 4758 | 15.80 |
| Anxiety | Any UI | 2011-12 | 1.08 (1.00, 1.18) | 0.06 | 3026 | 24.85 |
| Anxiety | Any Urgency | 2002-04 | 1.07 (0.97, 1.17) | 0.17 | 4758 | 10.59 |
| Anxiety | Any Urgency | 2011-12 | 1.08 (0.99, 1.19) | 0.10 | 3026 | 17.91 |
| Anxiety | Mixed UI | 2002-04 | 1.22 (1.05, 1.42) | 0.01 | 4758 | 3.93 |
| Anxiety | Mixed UI | 2011-12 | 1.02 (0.90, 1.17) | 0.71 | 3026 | 8.46 |
| Anxiety | Nocturia | 2002-04 | 1.08 (0.96, 1.22) | 0.20 | 4758 | 5.84 |
| Anxiety | Nocturia | 2011-12 | 1.12 (0.98, 1.27) | 0.09 | 3026 | 9.02 |
| Anxiety | Stress UI | 2002-04 | 1.05 (0.96, 1.16) | 0.28 | 4758 | 10.40 |
| Anxiety | Stress UI | 2011-12 | 1.10 (0.99, 1.22) | 0.08 | 3026 | 13.52 |
| Anxiety | Urgency UI | 2002-04 | 1.15 (0.90, 1.46) | 0.26 | 4758 | 1.47 |
| Anxiety | Urgency UI | 2011-12 | 1.08 (0.87, 1.34) | 0.48 | 3026 | 2.88 |
| Depression | Any UI | 2002-04 | 1.07 (0.99, 1.16) | 0.10 | 4758 | 15.80 |
| Depression | Any UI | 2011-12 | 1.08 (1.00, 1.18) | 0.06 | 3026 | 24.85 |
| Depression | Any Urgency | 2002-04 | 1.10 (1.00, 1.20) | 0.06 | 4758 | 10.59 |
| Depression | Any Urgency | 2011-12 | 1.13 (1.02, 1.24) | 0.01 | 3026 | 17.91 |
| Depression | Mixed UI | 2002-04 | 1.09 (0.94, 1.26) | 0.26 | 4758 | 3.93 |
| Depression | Mixed UI | 2011-12 | 1.02 (0.90, 1.16) | 0.74 | 3026 | 8.46 |
| Depression | Nocturia | 2002-04 | 1.30 (1.15, 1.47) | 0.00 | 4758 | 5.84 |
| Depression | Nocturia | 2011-12 | 1.24 (1.09, 1.40) | 0.00 | 3026 | 9.02 |
| Depression | Stress UI | 2002-04 | 1.05 (0.96, 1.16) | 0.27 | 4758 | 10.40 |
| Depression | Stress UI | 2011-12 | 1.06 (0.96, 1.18) | 0.26 | 3026 | 13.52 |
| Depression | Urgency UI | 2002-04 | 1.05 (0.83, 1.34) | 0.66 | 4758 | 1.47 |
| Depression | Urgency UI | 2011-12 | 1.23 (0.99, 1.53) | 0.06 | 3026 | 2.88 |
| Neuroticism | Any UI | 2002-04 | 1.13 (1.05, 1.22) | 0.00 | 4758 | 15.80 |
| Neuroticism | Any UI | 2011-12 | 1.03 (0.94, 1.11) | 0.54 | 3026 | 24.85 |
| Neuroticism | Any Urgency | 2002-04 | 1.09 (0.99, 1.19) | 0.07 | 4758 | 10.59 |
| Neuroticism | Any Urgency | 2011-12 | 1.06 (0.96, 1.16) | 0.25 | 3026 | 17.91 |
| Neuroticism | Mixed UI | 2002-04 | 1.22 (1.05, 1.41) | 0.01 | 4758 | 3.93 |
| Neuroticism | Mixed UI | 2011-12 | 0.97 (0.86, 1.11) | 0.68 | 3026 | 8.46 |
| Neuroticism | Nocturia | 2002-04 | 1.28 (1.13, 1.44) | 0.00 | 4758 | 5.84 |
| Neuroticism | Nocturia | 2011-12 | 1.16 (1.02, 1.31) | 0.02 | 3026 | 9.02 |
| Neuroticism | Stress UI | 2002-04 | 1.08 (0.99, 1.19) | 0.10 | 4758 | 10.40 |
| Neuroticism | Stress UI | 2011-12 | 1.04 (0.94, 1.16) | 0.44 | 3026 | 13.52 |
| Neuroticism | Urgency UI | 2002-04 | 1.10 (0.87, 1.40) | 0.41 | 4758 | 1.47 |
| Neuroticism | Urgency UI | 2011-12 | 1.08 (0.87, 1.34) | 0.49 | 3026 | 2.88 |

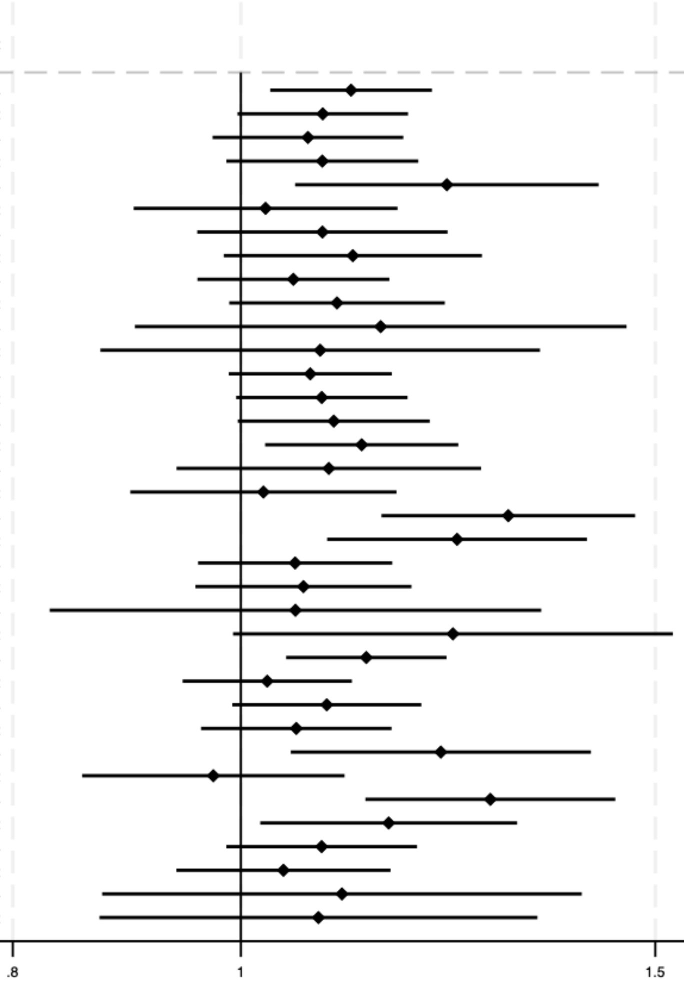

### Figure 6.7

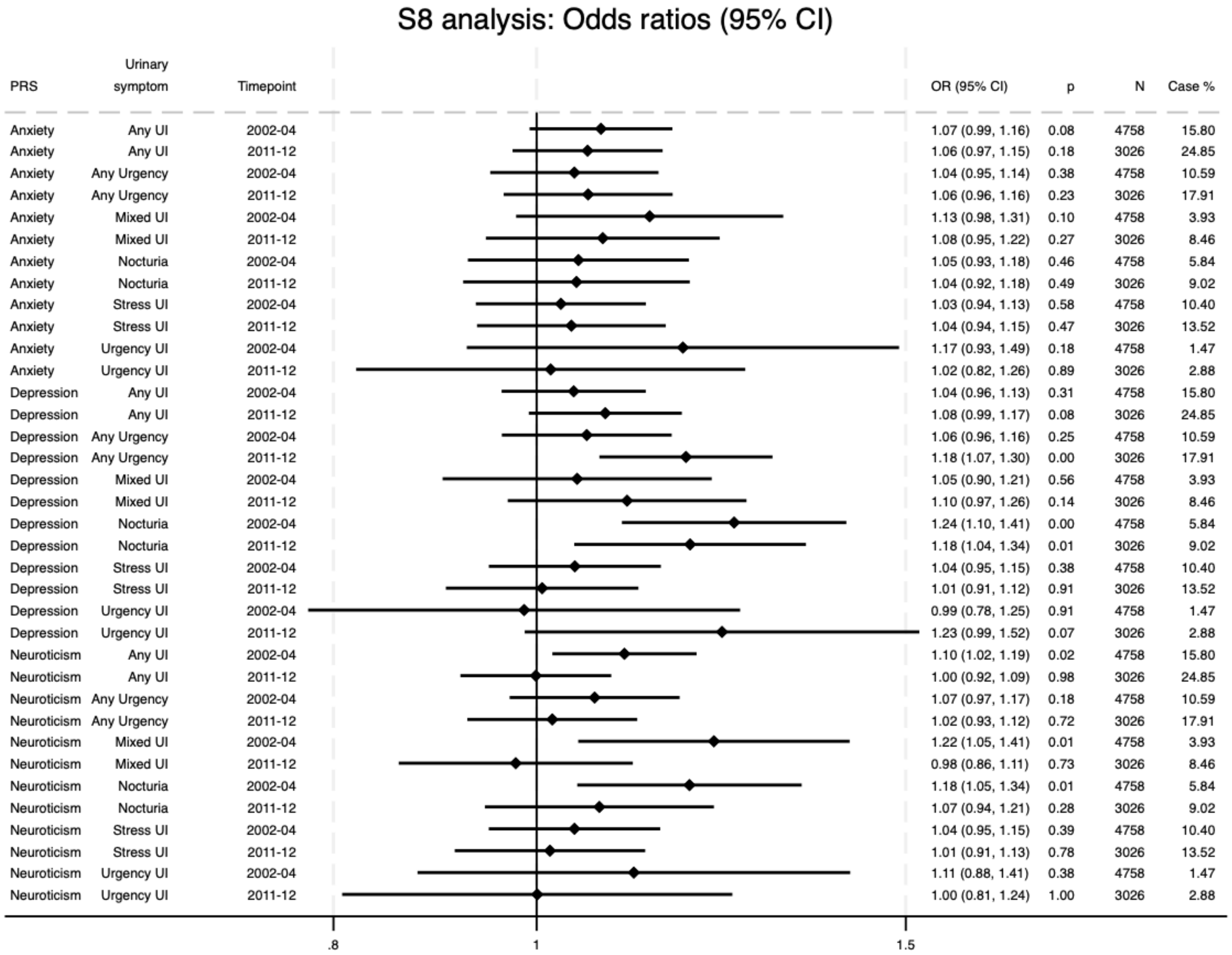
S8 analysis: Odds ratios (95% CI)
PRS
Urinary symptom
Timepoint
OR (95% CI)
p
N
Case %
Anxiety Any UI 2002-04 1.07 (0.99, 1.16) 0.08 4758 15.80
Anxiety Any UI 2011-12 1.06 (0.97, 1.15) 0.18 3026 24.85
Anxiety Any Urgency 2002-04 1.04 (0.95, 1.14) 0.38 4758 10.59
Anxiety Any Urgency 2011-12 1.06 (0.96, 1.16) 0.23 3026 17.91
Anxiety Mixed UI 2002-04 1.13 (0.98, 1.31) 0.10 4758 3.93
Anxiety Mixed UI 2011-12 1.08 (0.95, 1.22) 0.27 3026 8.46
Anxiety Nocturia 2002-04 1.05 (0.93, 1.18) 0.46 4758 5.84
Anxiety Nocturia 2011-12 1.04 (0.92, 1.18) 0.49 3026 9.02
Anxiety Stress UI 2002-04 1.03 (0.94, 1.13) 0.58 4758 10.40
Anxiety Stress UI 2011-12 1.04 (0.94, 1.15) 0.47 3026 13.52
Anxiety Urgency UI 2002-04 1.17 (0.93, 1.49) 0.18 4758 1.47
Anxiety Urgency UI 2011-12 1.02 (0.82, 1.26) 0.89 3026 2.88
Depression Any UI 2002-04 1.04 (0.96, 1.13) 0.31 4758 15.80
Depression Any UI 2011-12 1.08 (0.99, 1.17) 0.08 3026 24.85
Depression Any Urgency 2002-04 1.06 (0.96, 1.16) 0.25 4758 10.59
Depression Any Urgency 2011-12 1.18 (1.07, 1.30) 0.00 3026 17.91
Depression Mixed UI 2002-04 1.05 (0.90, 1.21) 0.56 4758 3.93
Depression Mixed UI 2011-12 1.10 (0.97, 1.26) 0.14 3026 8.46
Depression Nocturia 2002-04 1.24 (1.10, 1.41) 0.00 4758 5.84
Depression Nocturia 2011-12 1.18 (1.04, 1.34) 0.01 3026 9.02
Depression Stress UI 2002-04 1.04 (0.95, 1.15) 0.38 4758 10.40
Depression Stress UI 2011-12 1.01 (0.91, 1.12) 0.91 3026 13.52
Depression Urgency UI 2002-04 0.99 (0.78, 1.25) 0.91 4758 1.47
Depression Urgency UI 2011-12 1.23 (0.99, 1.52) 0.07 3026 2.88
Neuroticism Any UI 2002-04 1.10 (1.02, 1.19) 0.02 4758 15.80
Neuroticism Any UI 2011-12 1.00 (0.92, 1.09) 0.98 3026 24.85
Neuroticism Any Urgency 2002-04 1.07 (0.97, 1.17) 0.18 4758 10.59
Neuroticism Any Urgency 2011-12 1.02 (0.93, 1.12) 0.72 3026 17.91
Neuroticism Mixed UI 2002-04 1.22 (1.05, 1.41) 0.01 4758 3.93
Neuroticism Mixed UI 2011-12 0.98 (0.86, 1.11) 0.73 3026 8.46
Neuroticism Nocturia 2002-04 1.18 (1.05, 1.34) 0.01 4758 5.84
Neuroticism Nocturia 2011-12 1.07 (0.94, 1.21) 0.28 3026 9.02
Neuroticism Stress UI 2002-04 1.04 (0.95, 1.15) 0.39 4758 10.40
Neuroticism Stress UI 2011-12 1.01 (0.91, 1.13) 0.78 3026 13.52
Neuroticism Urgency UI 2002-04 1.11 (0.88, 1.41) 0.38 4758 1.47
Neuroticism Urgency UI 2011-12 1.00 (0.81, 1.24) 1.00 3026 2.88
.8
1
1.5

#### Figure 6.8

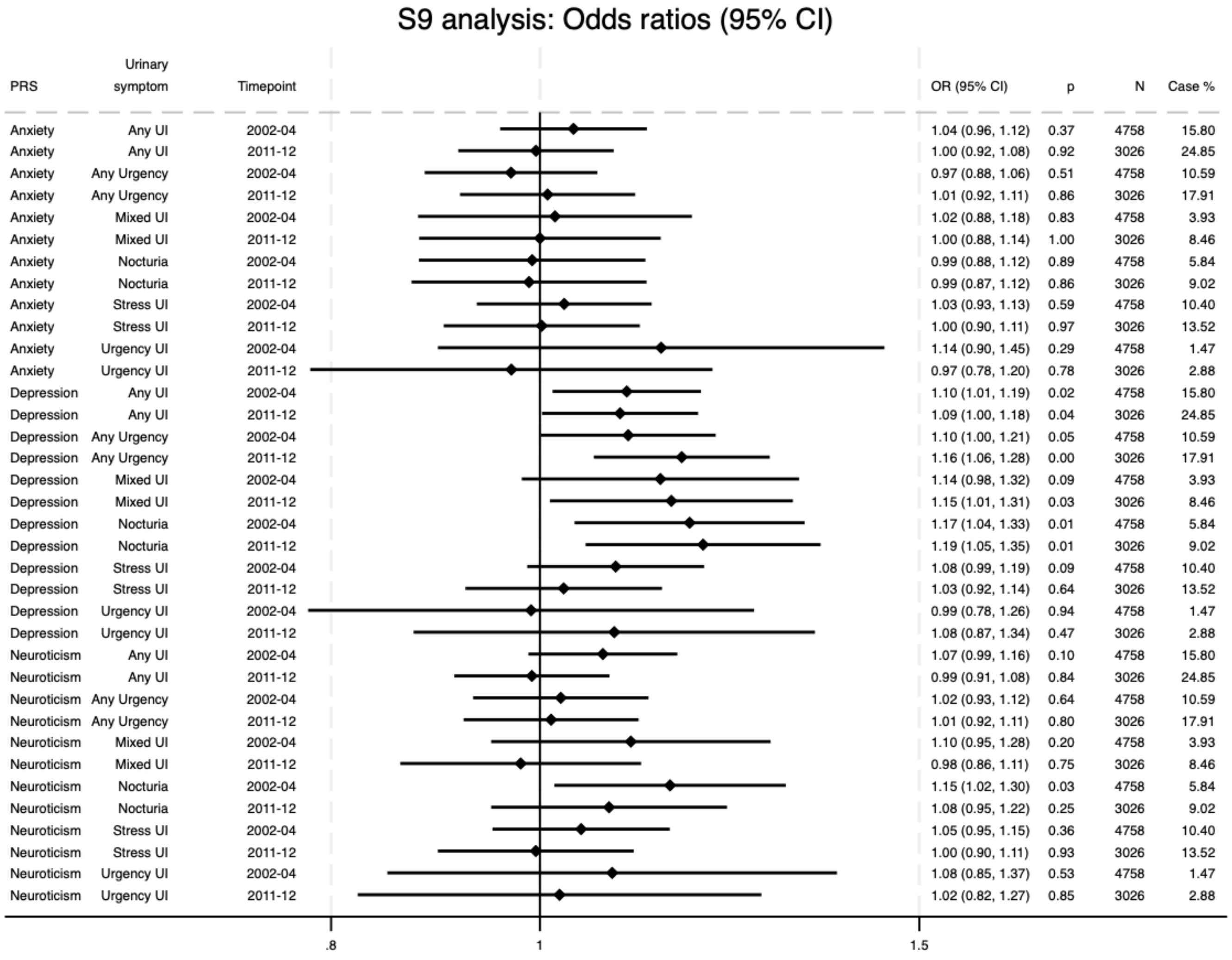
S9 analysis: Odds ratios (95% CI)
PRS Urinary symptom Timepoint OR (95% CI) p N Case %
Anxiety Any UI 2002-04 1.04 (0.96, 1.12) 0.37 4758 15.80
Anxiety Any UI 2011-12 1.00 (0.92, 1.08) 0.92 3026 24.85
Anxiety Any Urgency 2002-04 0.97 (0.88, 1.06) 0.51 4758 10.59
Anxiety Any Urgency 2011-12 1.01 (0.92, 1.11) 0.86 3026 17.91
Anxiety Mixed UI 2002-04 1.02 (0.88, 1.18) 0.83 4758 3.93
Anxiety Mixed UI 2011-12 1.00 (0.88, 1.14) 1.00 3026 8.46
Anxiety Nocturia 2002-04 0.99 (0.88, 1.12) 0.89 4758 5.84
Anxiety Nocturia 2011-12 0.99 (0.87, 1.12) 0.86 3026 9.02
Anxiety Stress UI 2002-04 1.03 (0.93, 1.13) 0.59 4758 10.40
Anxiety Stress UI 2011-12 1.00 (0.90, 1.11) 0.97 3026 13.52
Anxiety Urgency UI 2002-04 1.14 (0.90, 1.45) 0.29 4758 1.47
Anxiety Urgency UI 2011-12 0.97 (0.78, 1.20) 0.78 3026 2.88
Depression Any UI 2002-04 1.10 (1.01, 1.19) 0.02 4758 15.80
Depression Any UI 2011-12 1.09 (1.00, 1.18) 0.04 3026 24.85
Depression Any Urgency 2002-04 1.10 (1.00, 1.21) 0.05 4758 10.59
Depression Any Urgency 2011-12 1.16 (1.06, 1.28) 0.00 3026 17.91
Depression Mixed UI 2002-04 1.14 (0.98, 1.32) 0.09 4758 3.93
Depression Mixed UI 2011-12 1.15 (1.01, 1.31) 0.03 3026 8.46
Depression Nocturia 2002-04 1.17 (1.04, 1.33) 0.01 4758 5.84
Depression Nocturia 2011-12 1.19 (1.05, 1.35) 0.01 3026 9.02
Depression Stress UI 2002-04 1.08 (0.99, 1.19) 0.09 4758 10.40
Depression Stress UI 2011-12 1.03 (0.92, 1.14) 0.64 3026 13.52
Depression Urgency UI 2002-04 0.99 (0.78, 1.26) 0.94 4758 1.47
Depression Urgency UI 2011-12 1.08 (0.87, 1.34) 0.47 3026 2.88
Neuroticism Any UI 2002-04 1.07 (0.99, 1.16) 0.10 4758 15.80
Neuroticism Any UI 2011-12 0.99 (0.91, 1.08) 0.84 3026 24.85
Neuroticism Any Urgency 2002-04 1.02 (0.93, 1.12) 0.64 4758 10.59
Neuroticism Any Urgency 2011-12 1.01 (0.92, 1.11) 0.80 3026 17.91
Neuroticism Mixed UI 2002-04 1.10 (0.95, 1.28) 0.20 4758 3.93
Neuroticism Mixed UI 2011-12 0.98 (0.86, 1.11) 0.75 3026 8.46
Neuroticism Nocturia 2002-04 1.15 (1.02, 1.30) 0.03 4758 5.84
Neuroticism Nocturia 2011-12 1.08 (0.95, 1.22) 0.25 3026 9.02
Neuroticism Stress UI 2002-04 1.05 (0.95, 1.15) 0.36 4758 10.40
Neuroticism Stress UI 2011-12 1.00 (0.90, 1.11) 0.93 3026 13.52
Neuroticism Urgency UI 2002-04 1.08 (0.85, 1.37) 0.53 4758 1.47
Neuroticism Urgency UI 2011-12 1.02 (0.82, 1.27) 0.85 3026 2.88
.8
1
1.5

#### Figure 6.9

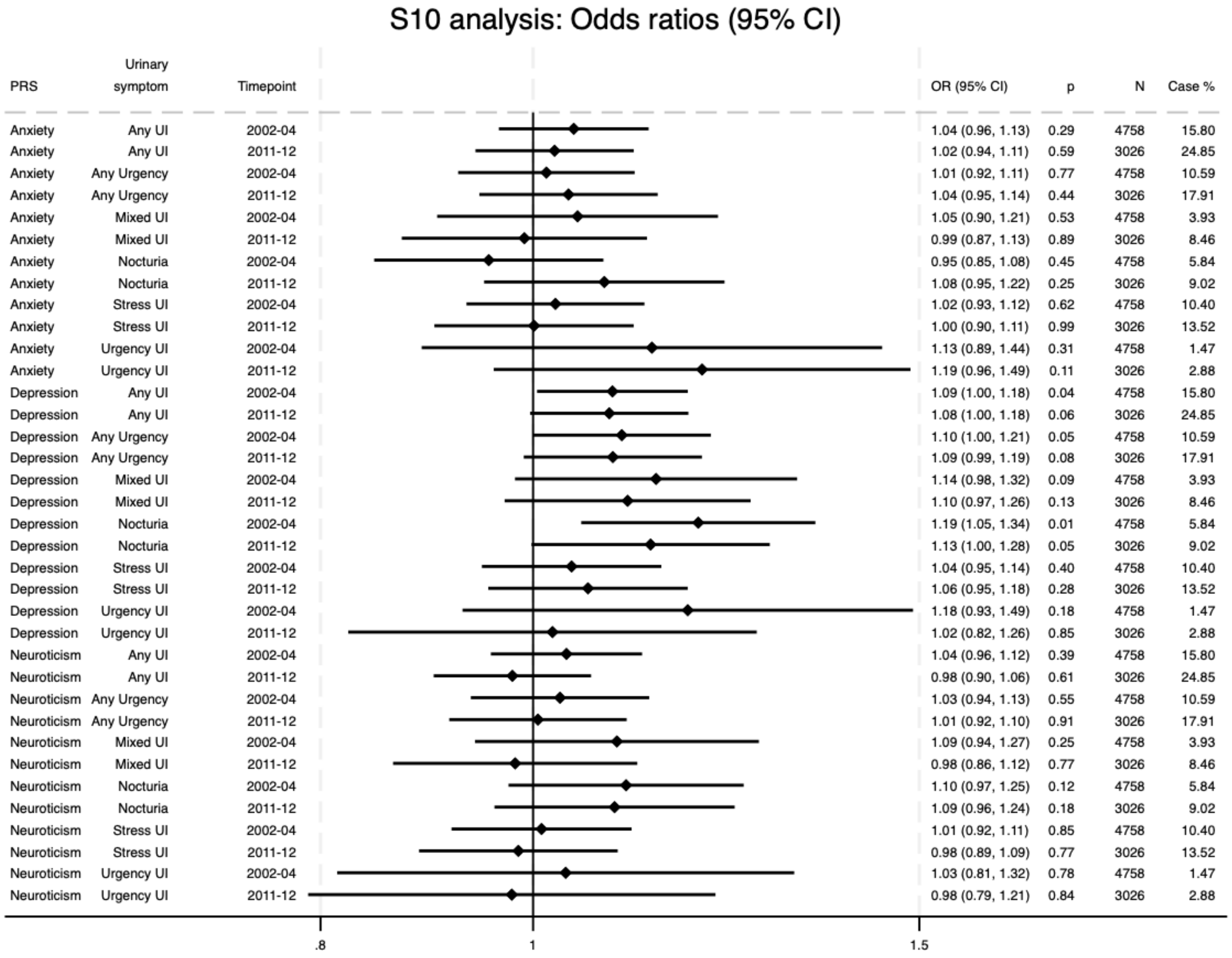
S10 analysis: Odds ratios (95% CI)
PRS | Urinary symptom | Timepoint | OR (95% CI) | p | N | Case %
Anxiety | Any UI | 2002-04 | 1.04 (0.96, 1.13) | 0.29 | 4758 | 15.80
Anxiety | Any UI | 2011-12 | 1.02 (0.94, 1.11) | 0.59 | 3026 | 24.85
Anxiety | Any Urgency | 2002-04 | 1.01 (0.92, 1.11) | 0.77 | 4758 | 10.59
Anxiety | Any Urgency | 2011-12 | 1.04 (0.95, 1.14) | 0.44 | 3026 | 17.91
Anxiety | Mixed UI | 2002-04 | 1.05 (0.90, 1.21) | 0.53 | 4758 | 3.93
Anxiety | Mixed UI | 2011-12 | 0.99 (0.87, 1.13) | 0.89 | 3026 | 8.46
Anxiety | Nocturia | 2002-04 | 0.95 (0.85, 1.08) | 0.45 | 4758 | 5.84
Anxiety | Nocturia | 2011-12 | 1.08 (0.95, 1.22) | 0.25 | 3026 | 9.02
Anxiety | Stress UI | 2002-04 | 1.02 (0.93, 1.12) | 0.62 | 4758 | 10.40
Anxiety | Stress UI | 2011-12 | 1.00 (0.90, 1.11) | 0.99 | 3026 | 13.52
Anxiety | Urgency UI | 2002-04 | 1.13 (0.89, 1.44) | 0.31 | 4758 | 1.47
Anxiety | Urgency UI | 2011-12 | 1.19 (0.96, 1.49) | 0.11 | 3026 | 2.88
Depression | Any UI | 2002-04 | 1.09 (1.00, 1.18) | 0.04 | 4758 | 15.80
Depression | Any UI | 2011-12 | 1.08 (1.00, 1.18) | 0.06 | 3026 | 24.85
Depression | Any Urgency | 2002-04 | 1.10 (1.00, 1.21) | 0.05 | 4758 | 10.59
Depression | Any Urgency | 2011-12 | 1.09 (0.99, 1.19) | 0.08 | 3026 | 17.91
Depression | Mixed UI | 2002-04 | 1.14 (0.98, 1.32) | 0.09 | 4758 | 3.93
Depression | Mixed UI | 2011-12 | 1.10 (0.97, 1.26) | 0.13 | 3026 | 8.46
Depression | Nocturia | 2002-04 | 1.19 (1.05, 1.34) | 0.01 | 4758 | 5.84
Depression | Nocturia | 2011-12 | 1.13 (1.00, 1.28) | 0.05 | 3026 | 9.02
Depression | Stress UI | 2002-04 | 1.04 (0.95, 1.14) | 0.40 | 4758 | 10.40
Depression | Stress UI | 2011-12 | 1.06 (0.95, 1.18) | 0.28 | 3026 | 13.52
Depression | Urgency UI | 2002-04 | 1.18 (0.93, 1.49) | 0.18 | 4758 | 1.47
Depression | Urgency UI | 2011-12 | 1.02 (0.82, 1.26) | 0.85 | 3026 | 2.88
Neuroticism | Any UI | 2002-04 | 1.04 (0.96, 1.12) | 0.39 | 4758 | 15.80
Neuroticism | Any UI | 2011-12 | 0.98 (0.90, 1.06) | 0.61 | 3026 | 24.85
Neuroticism | Any Urgency | 2002-04 | 1.03 (0.94, 1.13) | 0.55 | 4758 | 10.59
Neuroticism | Any Urgency | 2011-12 | 1.01 (0.92, 1.10) | 0.91 | 3026 | 17.91
Neuroticism | Mixed UI | 2002-04 | 1.09 (0.94, 1.27) | 0.25 | 4758 | 3.93
Neuroticism | Mixed UI | 2011-12 | 0.98 (0.86, 1.12) | 0.77 | 3026 | 8.46
Neuroticism | Nocturia | 2002-04 | 1.10 (0.97, 1.25) | 0.12 | 4758 | 5.84
Neuroticism | Nocturia | 2011-12 | 1.09 (0.96, 1.24) | 0.18 | 3026 | 9.02
Neuroticism | Stress UI | 2002-04 | 1.01 (0.92, 1.11) | 0.85 | 4758 | 10.40
Neuroticism | Stress UI | 2011-12 | 0.98 (0.89, 1.09) | 0.77 | 3026 | 13.52
Neuroticism | Urgency UI | 2002-04 | 1.03 (0.81, 1.32) | 0.78 | 4758 | 1.47
Neuroticism | Urgency UI | 2011-12 | 0.98 (0.79, 1.21) | 0.84 | 3026 | 2.88
.8
1
1.5

### Figure 6.10

#### S11 analysis: Odds ratios (95% CI)

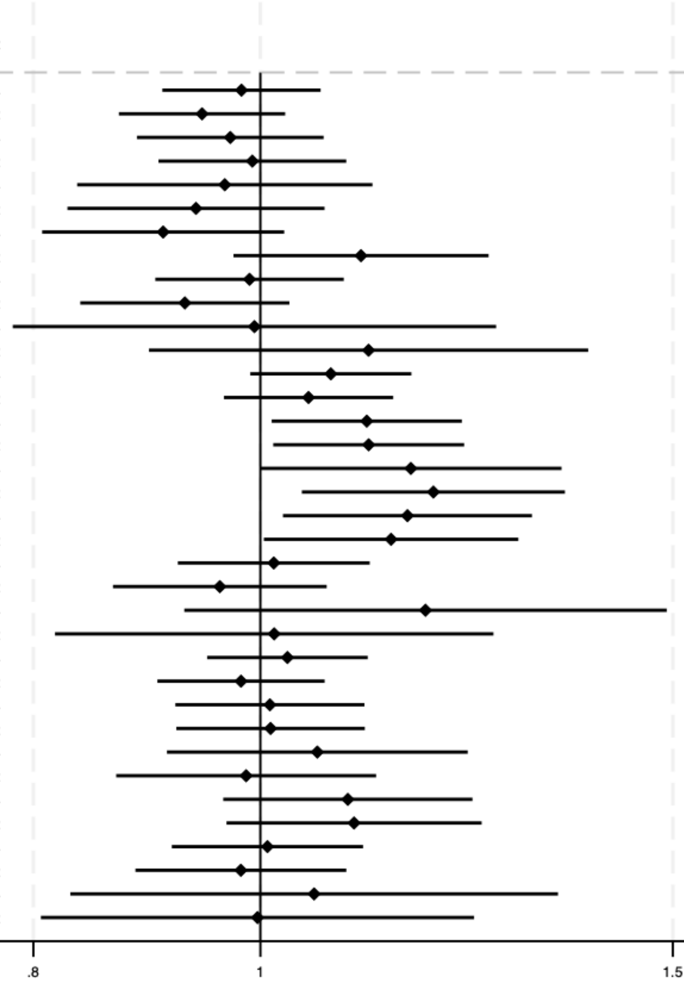

| PRS | Urinary symptom | Timepoint | OR (95% CI) | p | N | Case % |
|---|---|---|---|---|---|---|
| Anxiety | Any UI | 2002-04 | 0.98 (0.91, 1.06) | 0.64 | 4758 | 15.80 |
| Anxiety | Any UI | 2011-12 | 0.94 (0.87, 1.02) | 0.17 | 3026 | 24.85 |
| Anxiety | Any Urgency | 2002-04 | 0.97 (0.89, 1.06) | 0.53 | 4758 | 10.59 |
| Anxiety | Any Urgency | 2011-12 | 0.99 (0.90, 1.09) | 0.87 | 3026 | 17.91 |
| Anxiety | Mixed UI | 2002-04 | 0.97 (0.84, 1.12) | 0.64 | 4758 | 3.93 |
| Anxiety | Mixed UI | 2011-12 | 0.94 (0.83, 1.07) | 0.33 | 3026 | 8.46 |
| Anxiety | Nocturia | 2002-04 | 0.91 (0.81, 1.02) | 0.12 | 4758 | 5.84 |
| Anxiety | Nocturia | 2011-12 | 1.10 (0.97, 1.25) | 0.12 | 3026 | 9.02 |
| Anxiety | Stress UI | 2002-04 | 0.99 (0.90, 1.09) | 0.82 | 4758 | 10.40 |
| Anxiety | Stress UI | 2011-12 | 0.93 (0.84, 1.03) | 0.16 | 3026 | 13.52 |
| Anxiety | Urgency UI | 2002-04 | 0.99 (0.78, 1.26) | 0.96 | 4758 | 1.47 |
| Anxiety | Urgency UI | 2011-12 | 1.11 (0.90, 1.38) | 0.33 | 3026 | 2.88 |
| Depression | Any UI | 2002-04 | 1.07 (0.99, 1.16) | 0.09 | 4758 | 15.80 |
| Depression | Any UI | 2011-12 | 1.05 (0.96, 1.14) | 0.26 | 3026 | 24.85 |
| Depression | Any Urgency | 2002-04 | 1.11 (1.01, 1.22) | 0.03 | 4758 | 10.59 |
| Depression | Any Urgency | 2011-12 | 1.11 (1.01, 1.22) | 0.03 | 3026 | 17.91 |
| Depression | Mixed UI | 2002-04 | 1.16 (1.00, 1.34) | 0.05 | 4758 | 3.93 |
| Depression | Mixed UI | 2011-12 | 1.19 (1.04, 1.35) | 0.01 | 3026 | 8.46 |
| Depression | Nocturia | 2002-04 | 1.16 (1.02, 1.31) | 0.02 | 4758 | 5.84 |
| Depression | Nocturia | 2011-12 | 1.14 (1.00, 1.29) | 0.04 | 3026 | 9.02 |
| Depression | Stress UI | 2002-04 | 1.01 (0.92, 1.11) | 0.78 | 4758 | 10.40 |
| Depression | Stress UI | 2011-12 | 0.96 (0.87, 1.07) | 0.46 | 3026 | 13.52 |
| Depression | Urgency UI | 2002-04 | 1.18 (0.93, 1.49) | 0.18 | 4758 | 1.47 |
| Depression | Urgency UI | 2011-12 | 1.01 (0.82, 1.26) | 0.90 | 3026 | 2.88 |
| Neuroticism | Any UI | 2002-04 | 1.03 (0.95, 1.11) | 0.51 | 4758 | 15.80 |
| Neuroticism | Any UI | 2011-12 | 0.98 (0.90, 1.07) | 0.65 | 3026 | 24.85 |
| Neuroticism | Any Urgency | 2002-04 | 1.01 (0.92, 1.11) | 0.84 | 4758 | 10.59 |
| Neuroticism | Any Urgency | 2011-12 | 1.01 (0.92, 1.11) | 0.83 | 3026 | 17.91 |
| Neuroticism | Mixed UI | 2002-04 | 1.06 (0.91, 1.23) | 0.46 | 4758 | 3.93 |
| Neuroticism | Mixed UI | 2011-12 | 0.99 (0.87, 1.12) | 0.83 | 3026 | 8.46 |
| Neuroticism | Nocturia | 2002-04 | 1.09 (0.96, 1.23) | 0.17 | 4758 | 5.84 |
| Neuroticism | Nocturia | 2011-12 | 1.10 (0.97, 1.24) | 0.15 | 3026 | 9.02 |
| Neuroticism | Stress UI | 2002-04 | 1.01 (0.92, 1.11) | 0.88 | 4758 | 10.40 |
| Neuroticism | Stress UI | 2011-12 | 0.98 (0.88, 1.09) | 0.72 | 3026 | 13.52 |
| Neuroticism | Urgency UI | 2002-04 | 1.05 (0.83, 1.34) | 0.67 | 4758 | 1.47 |
| Neuroticism | Urgency UI | 2011-12 | 1.00 (0.81, 1.23) | 0.98 | 3026 | 2.88 |

#### Figure 6.11

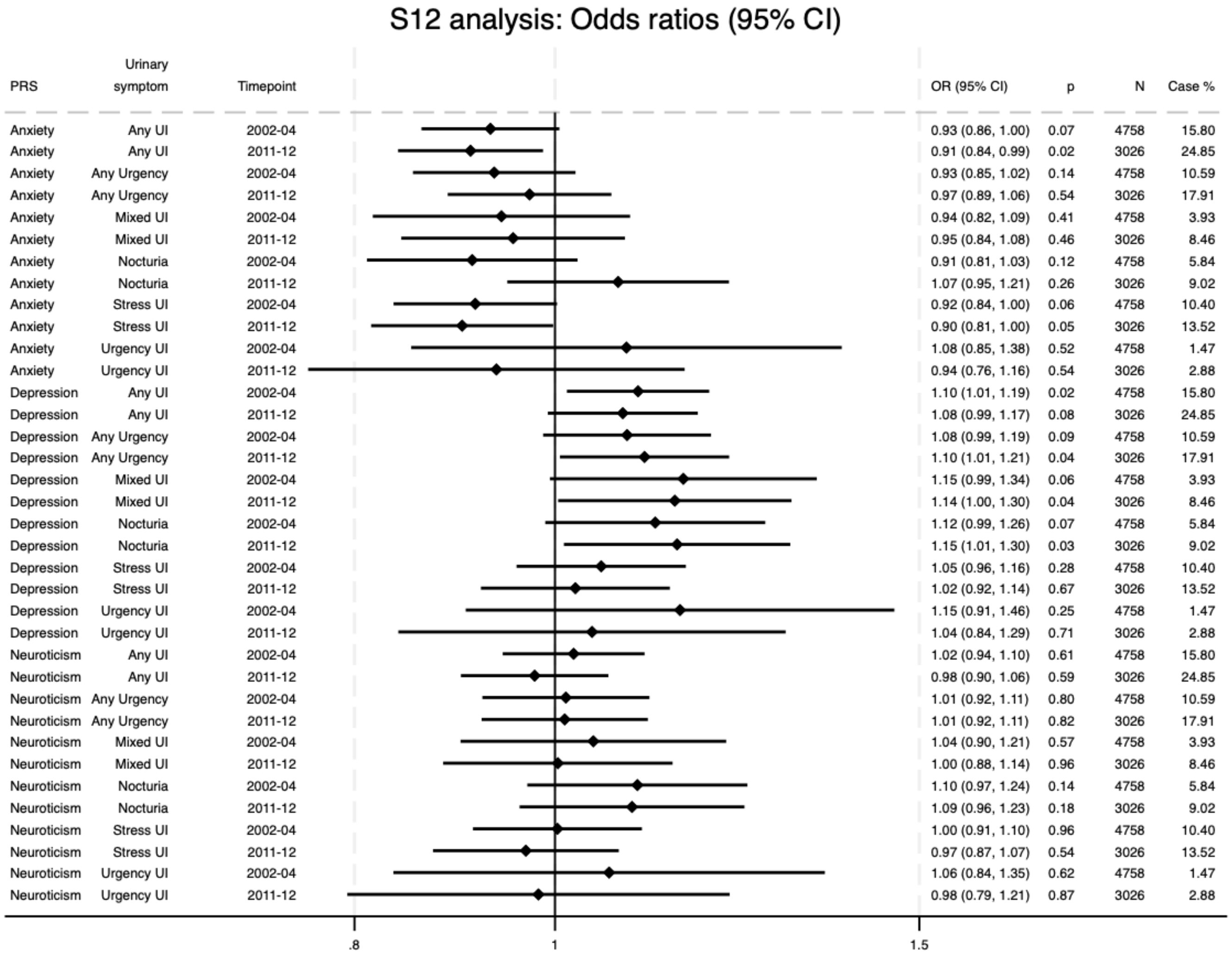
S12 analysis: Odds ratios (95% CI)
PRS | Urinary symptom | Timepoint | OR (95% CI) | p | N | Case %
Anxiety | Any UI | 2002-04 | 0.93 (0.86, 1.00) | 0.07 | 4758 | 15.80
Anxiety | Any UI | 2011-12 | 0.91 (0.84, 0.99) | 0.02 | 3026 | 24.85
Anxiety | Any Urgency | 2002-04 | 0.93 (0.85, 1.02) | 0.14 | 4758 | 10.59
Anxiety | Any Urgency | 2011-12 | 0.97 (0.89, 1.06) | 0.54 | 3026 | 17.91
Anxiety | Mixed UI | 2002-04 | 0.94 (0.82, 1.09) | 0.41 | 4758 | 3.93
Anxiety | Mixed UI | 2011-12 | 0.95 (0.84, 1.08) | 0.46 | 3026 | 8.46
Anxiety | Nocturia | 2002-04 | 0.91 (0.81, 1.03) | 0.12 | 4758 | 5.84
Anxiety | Nocturia | 2011-12 | 1.07 (0.95, 1.21) | 0.26 | 3026 | 9.02
Anxiety | Stress UI | 2002-04 | 0.92 (0.84, 1.00) | 0.06 | 4758 | 10.40
Anxiety | Stress UI | 2011-12 | 0.90 (0.81, 1.00) | 0.05 | 3026 | 13.52
Anxiety | Urgency UI | 2002-04 | 1.08 (0.85, 1.38) | 0.52 | 4758 | 1.47
Anxiety | Urgency UI | 2011-12 | 0.94 (0.76, 1.16) | 0.54 | 3026 | 2.88
Depression | Any UI | 2002-04 | 1.10 (1.01, 1.19) | 0.02 | 4758 | 15.80
Depression | Any UI | 2011-12 | 1.08 (0.99, 1.17) | 0.08 | 3026 | 24.85
Depression | Any Urgency | 2002-04 | 1.08 (0.99, 1.19) | 0.09 | 4758 | 10.59
Depression | Any Urgency | 2011-12 | 1.10 (1.01, 1.21) | 0.04 | 3026 | 17.91
Depression | Mixed UI | 2002-04 | 1.15 (0.99, 1.34) | 0.06 | 4758 | 3.93
Depression | Mixed UI | 2011-12 | 1.14 (1.00, 1.30) | 0.04 | 3026 | 8.46
Depression | Nocturia | 2002-04 | 1.12 (0.99, 1.26) | 0.07 | 4758 | 5.84
Depression | Nocturia | 2011-12 | 1.15 (1.01, 1.30) | 0.03 | 3026 | 9.02
Depression | Stress UI | 2002-04 | 1.05 (0.96, 1.16) | 0.28 | 4758 | 10.40
Depression | Stress UI | 2011-12 | 1.02 (0.92, 1.14) | 0.67 | 3026 | 13.52
Depression | Urgency UI | 2002-04 | 1.15 (0.91, 1.46) | 0.25 | 4758 | 1.47
Depression | Urgency UI | 2011-12 | 1.04 (0.84, 1.29) | 0.71 | 3026 | 2.88
Neuroticism | Any UI | 2002-04 | 1.02 (0.94, 1.10) | 0.61 | 4758 | 15.80
Neuroticism | Any UI | 2011-12 | 0.98 (0.90, 1.06) | 0.59 | 3026 | 24.85
Neuroticism | Any Urgency | 2002-04 | 1.01 (0.92, 1.11) | 0.80 | 4758 | 10.59
Neuroticism | Any Urgency | 2011-12 | 1.01 (0.92, 1.11) | 0.82 | 3026 | 17.91
Neuroticism | Mixed UI | 2002-04 | 1.04 (0.90, 1.21) | 0.57 | 4758 | 3.93
Neuroticism | Mixed UI | 2011-12 | 1.00 (0.88, 1.14) | 0.96 | 3026 | 8.46
Neuroticism | Nocturia | 2002-04 | 1.10 (0.97, 1.24) | 0.14 | 4758 | 5.84
Neuroticism | Nocturia | 2011-12 | 1.09 (0.96, 1.23) | 0.18 | 3026 | 9.02
Neuroticism | Stress UI | 2002-04 | 1.00 (0.91, 1.10) | 0.96 | 4758 | 10.40
Neuroticism | Stress UI | 2011-12 | 0.97 (0.87, 1.07) | 0.54 | 3026 | 13.52
Neuroticism | Urgency UI | 2002-04 | 1.06 (0.84, 1.35) | 0.62 | 4758 | 1.47
Neuroticism | Urgency UI | 2011-12 | 0.98 (0.79, 1.21) | 0.87 | 3026 | 2.88
.8
1
1.5

#### Figure 6.12

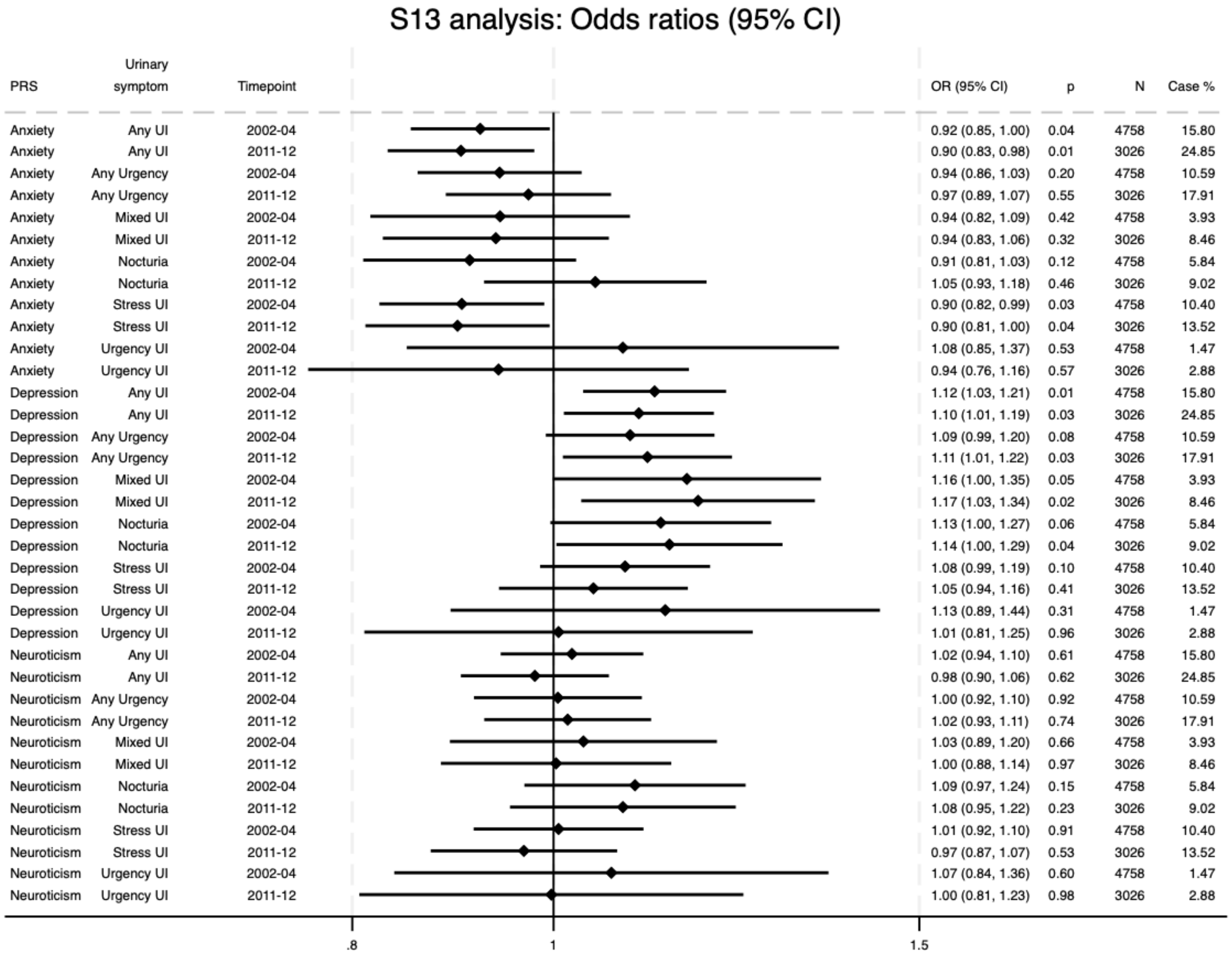
S13 analysis: Odds ratios (95% CI)
PRS
Urinary symptom
Timepoint
OR (95% CI)
p
N
Case %
Anxiety Any UI 2002-04 0.92 (0.85, 1.00) 0.04 4758 15.80
Anxiety Any UI 2011-12 0.90 (0.83, 0.98) 0.01 3026 24.85
Anxiety Any Urgency 2002-04 0.94 (0.86, 1.03) 0.20 4758 10.59
Anxiety Any Urgency 2011-12 0.97 (0.89, 1.07) 0.55 3026 17.91
Anxiety Mixed UI 2002-04 0.94 (0.82, 1.09) 0.42 4758 3.93
Anxiety Mixed UI 2011-12 0.94 (0.83, 1.06) 0.32 3026 8.46
Anxiety Nocturia 2002-04 0.91 (0.81, 1.03) 0.12 4758 5.84
Anxiety Nocturia 2011-12 1.05 (0.93, 1.18) 0.46 3026 9.02
Anxiety Stress UI 2002-04 0.90 (0.82, 0.99) 0.03 4758 10.40
Anxiety Stress UI 2011-12 0.90 (0.81, 1.00) 0.04 3026 13.52
Anxiety Urgency UI 2002-04 1.08 (0.85, 1.37) 0.53 4758 1.47
Anxiety Urgency UI 2011-12 0.94 (0.76, 1.16) 0.57 3026 2.88
Depression Any UI 2002-04 1.12 (1.03, 1.21) 0.01 4758 15.80
Depression Any UI 2011-12 1.10 (1.01, 1.19) 0.03 3026 24.85
Depression Any Urgency 2002-04 1.09 (0.99, 1.20) 0.08 4758 10.59
Depression Any Urgency 2011-12 1.11 (1.01, 1.22) 0.03 3026 17.91
Depression Mixed UI 2002-04 1.16 (1.00, 1.35) 0.05 4758 3.93
Depression Mixed UI 2011-12 1.17 (1.03, 1.34) 0.02 3026 8.46
Depression Nocturia 2002-04 1.13 (1.00, 1.27) 0.06 4758 5.84
Depression Nocturia 2011-12 1.14 (1.00, 1.29) 0.04 3026 9.02
Depression Stress UI 2002-04 1.08 (0.99, 1.19) 0.10 4758 10.40
Depression Stress UI 2011-12 1.05 (0.94, 1.16) 0.41 3026 13.52
Depression Urgency UI 2002-04 1.13 (0.89, 1.44) 0.31 4758 1.47
Depression Urgency UI 2011-12 1.01 (0.81, 1.25) 0.96 3026 2.88
Neuroticism Any UI 2002-04 1.02 (0.94, 1.10) 0.61 4758 15.80
Neuroticism Any UI 2011-12 0.98 (0.90, 1.06) 0.62 3026 24.85
Neuroticism Any Urgency 2002-04 1.00 (0.92, 1.10) 0.92 4758 10.59
Neuroticism Any Urgency 2011-12 1.02 (0.93, 1.11) 0.74 3026 17.91
Neuroticism Mixed UI 2002-04 1.03 (0.89, 1.20) 0.66 4758 3.93
Neuroticism Mixed UI 2011-12 1.00 (0.88, 1.14) 0.97 3026 8.46
Neuroticism Nocturia 2002-04 1.09 (0.97, 1.24) 0.15 4758 5.84
Neuroticism Nocturia 2011-12 1.08 (0.95, 1.22) 0.23 3026 9.02
Neuroticism Stress UI 2002-04 1.01 (0.92, 1.10) 0.91 4758 10.40
Neuroticism Stress UI 2011-12 0.97 (0.87, 1.07) 0.53 3026 13.52
Neuroticism Urgency UI 2002-04 1.07 (0.84, 1.36) 0.60 4758 1.47
Neuroticism Urgency UI 2011-12 1.00 (0.81, 1.23) 0.98 3026 2.88
.8
1
1.5
